# Spatial Structure of Tumor and Immune Cells Shape Outcomes in ER⁺HER2⁻ and Triple-Negative Breast Cancer

**DOI:** 10.1101/2025.11.02.25338835

**Authors:** Iris Nederlof, Siamak Hajizadeh, Rolf Harkes, Barbara Andrade Barbosa, Michiel de Maaker, Ingrid Hofland, Dennis Peters, Robert Elens, Annegien Broeks, Erik van Werkhoven, Yongsoo Kim, Marieke Ijsselstein, Noel F.C.C. de Miranda, Sophie Bosma, Nicole C. M. Visser, Malou L.H. Snijders, Joyce Sanders, Roberto Salgado, Philip Poortmans, Astrid Scholten, Liesbeth J. Boersma, Marleen Kok, Harry Bartelink, Marc J. Van De Vijver, Hugo M. Horlings

## Abstract

Immune infiltration is prognostic in triple-negative breast cancer (TNBC), but its role in ER⁺/HER2⁻ disease remains unclear, and conventional scoring may overlook spatial context. We analyzed tumors from 1,037 women ≤50 years in the Young Boost Trial (NCT00212121), integrating centralized pathology review, deep learning–based spatial profiling of whole-slide H&E, and multiplex immunophenotyping.

In TNBC, stromal tumor-infiltrating lymphocytes (sTILs) were robustly prognostic, independent of clinicopathologic features, confirming immune burden as the dominant signal. By contrast, in ER⁺/HER2⁻ cancers, sTILs were protective only in low-grade tumors and lost significance when architectural features were considered. Instead, grade, lymphovascular invasion, central sclerosis, and spatial separation of lymphocytes and tumor cells carried stronger independent prognostic value.

These findings highlight subtype-specific prognostic biology: in TNBC, immune density alone captures outcome, whereas in ER⁺/HER2⁻ disease, the interplay between immune infiltrates and tumor architecture governs prognostic associations.

## Introduction

Immune infiltration in breast cancer is highly variable and can strongly influence clinical outcomes.^1–3^ Among established biomarkers, stromal tumor-infiltrating lymphocytes (sTILs) are consistently prognostic in triple-negative breast cancer (TNBC), but their utility in ER⁺HER2⁻ disease is less clear, with associations often weak or inconsistent.^4,5^ Beyond immune quantity, increasing evidence suggests that spatial organization, such as exclusion, clustering, or mixing with tumor cells, also shapes anti-tumor immunity.^6–9^

Recently, two large randomized phase III trials demonstrated that adding immune checkpoint inhibitors to neoadjuvant chemotherapy improves pathologic complete response rates in high-risk, early-stage ER⁺HER2⁻ breast cancer ^10,11^. These results highlight the therapeutic potential of immunotherapy in luminal breast cancers and reinforce the importance of understanding how immune contexture, including both abundance and spatial architecture, modulates disease biology.

Immune patterns, however, do not emerge in isolation; they are shaped by the structural constraints of the tumor microenvironment. Architectural features such as stromal density, fibrosis, necrosis, and border morphology regulate immune accessibility and function.^12–16^ In ER⁺HER2⁻ cancers, where immune infiltration is typically sparse ^3^, architectural variability may mask or modulate immune signals^17–20^. In contrast, in TNBC, the immune burden itself often dominates. Classical pathology, including tumor grade, stromal composition, and necrosis,^21,22^ already captures some aspects of this ecology, yet systematic integration of architectural and immune spatial metrics remains limited.

Most current profiling efforts prioritize either immune abundance or molecular correlates, often overlooking the architectural context in which immune cells are embedded.^23–26^ Structural features such as fibrosis and border morphology do not merely mirror tumor aggressiveness; they actively shape immune topography and retention within the microenvironment ^8,9,14,19^. A clearer understanding of how immune and structural features interact is needed to refine risk stratification and identify subtype-specific prognostic markers.

Here, we leveraged a large prospective cohort from the Young Boost Trial (NCT00212121), comprising 1,037 primary breast cancers, including 797 ER⁺HER2⁻ tumors, with long clinical follow-up and centralized pathology review. Using deeplearning–based spatial profiling of routine H&E slides alongside multiplex immunophenotyping, we systematically quantified both immune infiltration and tumor architecture. This unique setting, free from confounding effects of neoadjuvant chemotherapy or immunotherapy, provides an opportunity to decode the tumor microenvironment of ER⁺HER2⁻ breast cancer in its untreated state. We demonstrate that prognostic signals are context dependent: immune burden is dominant in TNBC, whereas in ER⁺HER2⁻ cancers, architectural features and immune–tumor separation carry greater independent weight.

## Results

### Cohort and endpoint

We analyzed tumor specimens from patients with primary invasive breast cancer without distant metastases at diagnosis (M0), enrolled in a prospective, multicenter study (Young Boost Trial; NCT00212121) across the Netherlands, France, and Germany (2004–2011)^27,28^. Eligible patients were ≤50 years. Of 2,421 participants, archival FFPE tissue was retrieved for 1,481, and a representative, evaluable tumor block was available for 1,370 (57%). Baseline clinical characteristics were comparable between patients with available tissue (n=1,370) and those without (n=1,051; **Supplementary Table 1**). Among evaluable tumors, 797 were ER⁺HER2⁻ and 240 were triple-negative (TNBC; ER ≤10%^29^, per national guidelines). Most patients presented with relatively early-stage disease: 69.8% had T1 tumors and 70% were node-negative (**Supplementary Table 1**). Over a median follow-up of 89 months, 168 patients developed distant metastases (M+) and 194 died. The primary endpoint was distant metastasis–free survival (DMFS).

### Centralized tumor architectural and immune evaluation

Whole-slide H&E sections from 1,370 evaluable tumors underwent centralized review on an online pathology platform (SlideScore^30^; results in **Table 1**). Pathologists scored histological subtype, grade, tumor border configuration, necrosis, fibrosis^31–36^ and sTILs^5^ (**Table 2**). While sTILs are an established biomarker in breast cancer, other immune features remain less characterized. We initially sought to assess immune exclusion, but this phenotype was identified in only three TNBC cases, underscoring that H&E review does not recapitulate exclusion patterns previously reported using CD8 immunohistochemistry ^8,9^. This prompted us to extend analyses with computational pathology.

**Table 1.** Clinicopathological and immune features in estrogen receptor-positive/HER2-negative (ER⁺HER2⁻) and triple-negative breast cancer (TNBC) tumors, stratified by distant metastatic outcome (M− vs. M+). HER2⁺ tumors were excluded. Continuous variables (e.g., age, sTILs) are presented as mean (standard deviation) and range; categorical variables are reported as counts and percentages. Comparisons were made using the Kruskal–Wallis rank sum test for continuous variables and Pearson’s chi-squared test for categorical variables. Bold p-values indicate statistical significance (p < 0.05).

| Feature | ER<sup>+</sup>HER2<sup>-</sup> M<sup>-</sup> (N = 738) | ER<sup>+</sup>HER2<sup>-</sup> M<sup>+</sup> (N = 59) | p | TNBC M<sup>-</sup> (N = 206) | TNBC M<sup>+</sup> (N = 34) | p |
| --- | --- | --- | --- | --- | --- | --- |
| Age (years) | 44.8 (4.4) | 45.5 (3.4) | 0.461 | 41.1 (6.2) | 41.5 (6.0) | 0.724 |
| Range | 27–51 | 36–50 |  | 24–50 | 30–50 |  |
| Menopausal status |  |  | 0.845 |  |  | 0.693 |
| Post | 56 (7.9%) | 5 (8.6%) |  | 14 (6.9%) | 3 (8.8%) |  |
| Pre | 653 (92.1%) | 53 (91.4%) |  | 188 (93.1%) | 31 (91.2%) |  |
| Tumor size |  |  | 0.049 |  |  | 0.460 |
| T1 | 558 (75.9%) | 38 (64.4%) |  | 117 (56.8%) | 17 (50.0%) |  |
| T2–3 | 177 (24.1%) | 21 (35.6%) |  | 89 (43.2%) | 17 (50.0%) |  |
| Nodal status |  |  | 0.042 |  |  | 0.735 |
| N0 | 518 (70.3%) | 34 (57.6%) |  | 157 (76.2%) | 25 (73.5%) |  |
| N+ | 219 (29.7%) | 25 (42.4%) |  | 49 (23.8%) | 9 (26.5%) |  |
| Histological subtype |  |  | 0.726 |  |  | 0.462 |
| Lobular | 53 (7.2%) | 3 (5.1%) |  | 2 (1.0%) | 0 (0.0%) |  |
| NST | 658 (89.2%) | 53 (89.8%) |  | 197 (95.6%) | 34 (100.0%) |  |
| Other | 27 (3.7%) | 3 (5.1%) |  | 7 (3.4%) | 0 (0.0%) |  |
| Tumor grade |  |  | 0.054 |  |  | 0.741 |
| 1 | 172 (23.3%) | 6 (10.2%) |  | 6 (2.9%) | 1 (2.9%) |  |
| 2 | 394 (53.4%) | 39 (66.1%) |  | 37 (18.0%) | 8 (23.5%) |  |
| 3 | 172 (23.3%) | 14 (23.7%) |  | 163 (79.1%) | 25 (73.5%) |  |
| sTILs (%) |  |  | 0.049 |  |  | <0.001 |
| Mean (SD) | 11.5 (14.3) | 8.6 (11.1) |  | 35.4 (26.0) | 19.2 (16.8) |  |
| Range | 0–80 | 0–60 |  | 1–90 | 1–80 |  |
| Stroma in tumor (%) |  |  | 0.698 |  |  | 0.118 |
| Mean (SD) | 41.6 (21.6) | 42.7 (22.9) |  | 31.4 (23.2) | 36.7 (20.3) |  |
| Range | 1–95 | 5–80 |  | 1–90 | 2–70 |  |
| Any necrosis |  |  | 0.640 |  |  | 0.521 |
| Absent | 485 (65.7%) | 37 (62.7%) |  | 53 (25.7%) | 7 (20.6%) |  |
| Present | 253 (34.3%) | 22 (37.3%) |  | 153 (74.3%) | 27 (79.4%) |  |
| Tumor border |  |  | 0.734 |  |  | 0.559 |
| Infiltrative | 490 (66.6%) | 38 (64.4%) |  | 45 (22.0%) | 9 (26.5%) |  |
| Pushing | 246 (33.4%) | 21 (35.6%) |  | 160 (78.0%) | 25 (73.5%) |  |
| Central sclerosis |  |  | 0.010 |  |  | 0.125 |
| Absent | 452 (61.2%) | 26 (44.1%) |  | 114 (55.3%) | 14 (41.2%) |  |
| Present | 286 (38.8%) | 33 (55.9%) |  | 92 (44.7%) | 20 (58.8%) |  |
| Lymphovascular invasion |  |  | 0.022 |  |  | 0.003 |
| Absent | 669 (90.7%) | 48 (81.4%) |  | 193 (94.1%) | 27 (79.4%) |  |
| Present | 69 (9.3%) | 11 (18.6%) |  | 12 (5.9%) | 7 (20.6%) |  |
| Immune hotspots |  |  | 0.050 |  |  | 0.254 |
| No hotspots | 497 (67.3%) | 47 (79.7%) |  | 100 (48.8%) | 13 (38.2%) |  |
| Hotspots (1 or more) | 241 (32.7%) | 12 (20.3%) |  | 105 (51.2%) | 21 (61.8%) |  |
| Immune phenotype excluded |  |  | 0.665 |  |  | 0.009 |
| Excluded | 29 (3.9%) | 3 (5.1%) |  | 1 (0.5%) | 2 (5.9%) |  |
| Not excluded | 708 (96.1%) | 56 (94.9%) |  | 204 (99.5%) | 32 (94.1%) |  |
| In situ component |  |  | 0.311 |  |  | 0.096 |
| No | 311 (42.4%) | 29 (49.2%) |  | 146 (71.9%) | 19 (57.6%) |  |
| Yes | 423 (57.6%) | 30 (50.8%) |  | 57 (28.1%) | 14 (42.4%) |  |

**Abbreviations:** ER, estrogen receptor; HER2, human epidermal growth factor receptor 2; TNBC, triple-negative breast cancer; M-, metastasis-negative; M+, metastasis-positive; sTILs, stromal tumor-infiltrating lymphocytes; NST, no special type (invasive ductal carcinoma); LVI, lymphovascular invasion.

**Table 2.** Overview of Tumor Architecture and Immune Scoring Features. Summary of the histological and computational features used to characterize tumor architecture and immune infiltration (see Methods for detailed description). Whole-slide H&E-stained sections were assessed by six expert breast pathologists using SlideScore^30^. Pathology-based features (e.g., tumor grade, stroma ratio, necrosis) were scored per international guidelines. Stromal tumor-infiltrating lymphocytes (sTILs) were quantified on a continuous scale (0–100%) following the 2015 international working group guidelines. Additional spatial and cellular metrics were derived using a deep learning model trained on cohort-specific annotations. The model enabled cell classification (tumor, lymphocyte, other) and spatial profiling to calculate immune proximity and exclusion (e.g., KL divergence and iTIL). The intratumoral TIL (iTIL) metric was defined as: *iTIL = lymphocyte fraction × (−normalized tumor→lymphocyte KL divergence)* Higher iTIL reflects greater immune burden with enhanced tumor–immune co-localization.

| Feature | Scoring / Scale | Clinical Significance | Routine Reporting | Assessed By |
| --- | --- | --- | --- | --- |
| Histology | NST, lobular, cribriform, metaplastic, etc. | Influences prognosis and treatment | Yes | Pathologists |
| Tumor Grade | Grade 1–3 (Bloom–Richardson) | Reflects tumor aggressiveness | Yes | Pathologists |
| Tumor Border | % pushing vs. infiltrative (0–100%) | Linked to invasion and prognosis | Yes | Pathologists |
| sTILs | 0–100% stromal lymphocytes | Prognostic in TNBC and ER<sup>+</sup> tumors | Yes | Pathologists |
| Central Fibrosis / Sclerosis | Present / absent | Indicates stromal remodeling, immune exclusion | Yes | Pathologists |
| Tumor Necrosis | Present / absent | Associated with aggressive biology | Yes | Pathologists |
| Stroma Ratio (SR) | <50% / ≥50% | Prognostic across cancers | Occasionally | Pathologists |
| Model derived lymphocyte density | % of lymphocytes (among all cells) | Quantitative; correlates with sTILs | No | Computational |
| Model derived tumor density | % of tumor cells (among all cells) | Reflects tumor burden | No | Computational |
| Tumor → Nearest Lymphocyte | Mean/min distance | Indicates immune accessibility to tumor | No | Computational |
| Lymphocyte → Nearest Tumor | Mean/min distance | Complements infiltration directionality | No | Computational |
| KL Divergence | Continuous score | Measures immune–tumor spatial separation | No | Computational |
| iTIL | Composite metric | Integrates abundance and proximity | No | Computational |

**Abbreviations:** NST, No Special Type; sTILs, Stromal Tumor-Infiltrating Lymphocytes; SR, Stroma Ratio; KL, Kullback–Leibler; TLS, Tertiary Lymphoid Structure; iTIL, intratumoral TIL score; AI, Artificial Intelligence.

To standardize single-cell spatial profiling of H&E slides, we developed a deep-learning pipeline trained on cohort-specific data (**Fig. 1a–c**). The model achieved high performance for tumor and immune-cell classification (balanced accuracy 0.958; **Fig. 1b–c**) and generated standardized spatial features, including cell-type fractions/densities, bidirectional nearest-neighbor distances (tumor→lymphocyte and lymphocyte→tumor), and Kullback–Leibler (KL) divergence to quantify immune–tumor separation (**Table 2**). In addition, we defined a composite *iTIL* metric integrating immune abundance and mixing: iTIL = lymphocyte fraction × (−normalized tumor→lymphocyte KL divergence), where higher values reflect greater immune burden with closer tumor–immune co-localization.

**Figure 1.**
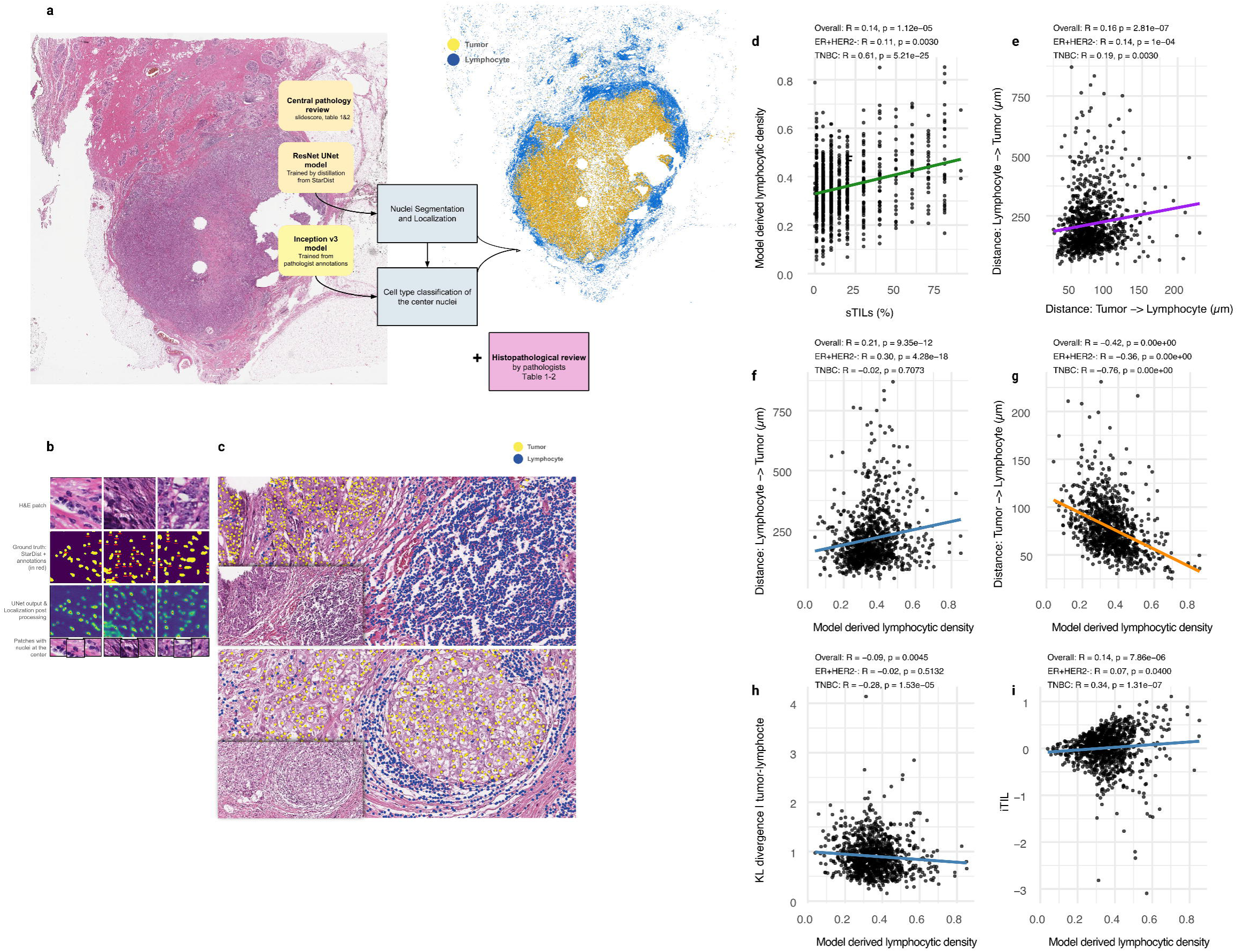
| H&E-derived immune quantification and spatial organization in ER+HER2- and TNBC. a–c, Representative H&E fields and computational workflow. A convolutional pipeline (nuclei segmentation with a ResNet–U-Net distilled from StarDist; cell-type classification with an DenseNet 161 model trained by breast pathologists) detects nuclei and assigns cell identity. Overlays show tumor cells (yellow) and lymphocytes (blue); inset, whole-slide overview of the same case. All H&E whole-slide images were also centrally reviewed and scored by breast pathologists for histopathologic features (including sTILs) according to guidelines; Table 1-2. d, Concordance of deep-learning (DL) lymphocyte ratio (lymphocytes / all cells) with pathologist-scored sTILs; points colored by subtype with Spearman’s ρ and two-sided p values. e, distance tumor --> lymphocyte vs lymphocyte -> tumor; points colored by subtype with Spearman’s ρ and two-sided p values. f-i, DL lymphocyte ratio versus spatial proximity metrics: (f) lymphocyte to tumor distance; (g) tumor to lymphocyte distance; (h) KL divergence; i iTIL; Spearman’s ρ and p values shown. Scatter plots show least-squares fit with 95% CI. Scale bars as shown.

Model-derived lymphocyte density correlated with pathologist-scored sTILs (**Fig. 1d**), with stronger concordance in TNBC (R = 0.61) than in ER⁺HER2⁻ (R=0.11). The weaker correlation in ER⁺HER2⁻ likely reflects pathologists’ implicit weighting of spatial context (e.g., clustering at tumor–stroma interfaces), whereas the model quantifies global density across the slide.

Bidirectional nearest-neighbor distances—tumor→lymphocyte and lymphocyte→tumor—were only weakly correlated with each other (R=0.16; **Fig. 1e**), indicating that they capture distinct spatial signals. In TNBC, increasing lymphocyte density was not associated with changes in lymphocyte→tumor distance (R = -0.02), suggesting stable infiltration patterns. By contrast, in ER⁺HER2⁻ tumors, higher lymphocyte density was accompanied by greater lymphocyte→tumor distance (R=0.31; **Fig. 1f**), consistent with peripheral rather than intermingled localization. Conversely, tumor→lymphocyte distance decreased with rising lymphocyte density in both TNBC and ER⁺HER2⁻ (R=-0.42; **Fig. 1g**), reflecting closer tumor proximity as immune burden increased.

Consistently, in TNBC lymphocyte density was inversely related to KL divergence (a measure of tumor–lymphocyte separation) (R=−0.27; **Fig. 1h**) and positively related to iTIL (R=0.34; **Fig. 1i**), whereas in ER⁺HER2⁻ this coupling was absent (R=−0.02 and 0.04; **Fig. 1h,i**), indicating that in luminal cancers immune abundance does not necessarily translate into closer tumor–immune apposition.

### Divergent prognostic roles of immune quantity and spatial architecture by subtype

As expected, sTILs were higher in TNBC than ER⁺HER2⁻ (**Supplementary Fig. 1a**). Given distinct clinical implications, analyses were stratified by subtype. In both cohorts, patients who developed distant metastases (M⁺) had lower baseline sTILs (**Table 1**; **Supplementary Fig. 1b-c**). Kaplan–Meier curves showed borderline separation in ER⁺HER2⁻ at >5% vs ≤5% sTILs (HR 0.60; 95% CI 0.36–1.00; p=0.054; **Fig. 2a**) and significant separation in TNBC at ≥50% versus <50% sTILs (p=0.012; **Fig. 2b**). In univariate analyses, sTILs were protective in TNBC (HR 0.80 per 10%; 95% CI 0.68–0.94; **Fig. 2g**) and among chemotherapy-treated patients (HR 0.81; 95% CI 0.69–0.95; **Fig. S1f**). In ER⁺HER2⁻ disease, associations were weaker overall (**Fig. 2h**) but reached significance in grade 1–2 tumors (HR 0.51; 95% CI 0.29–0.89; **Fig. S1d**).

**Figure 2.**
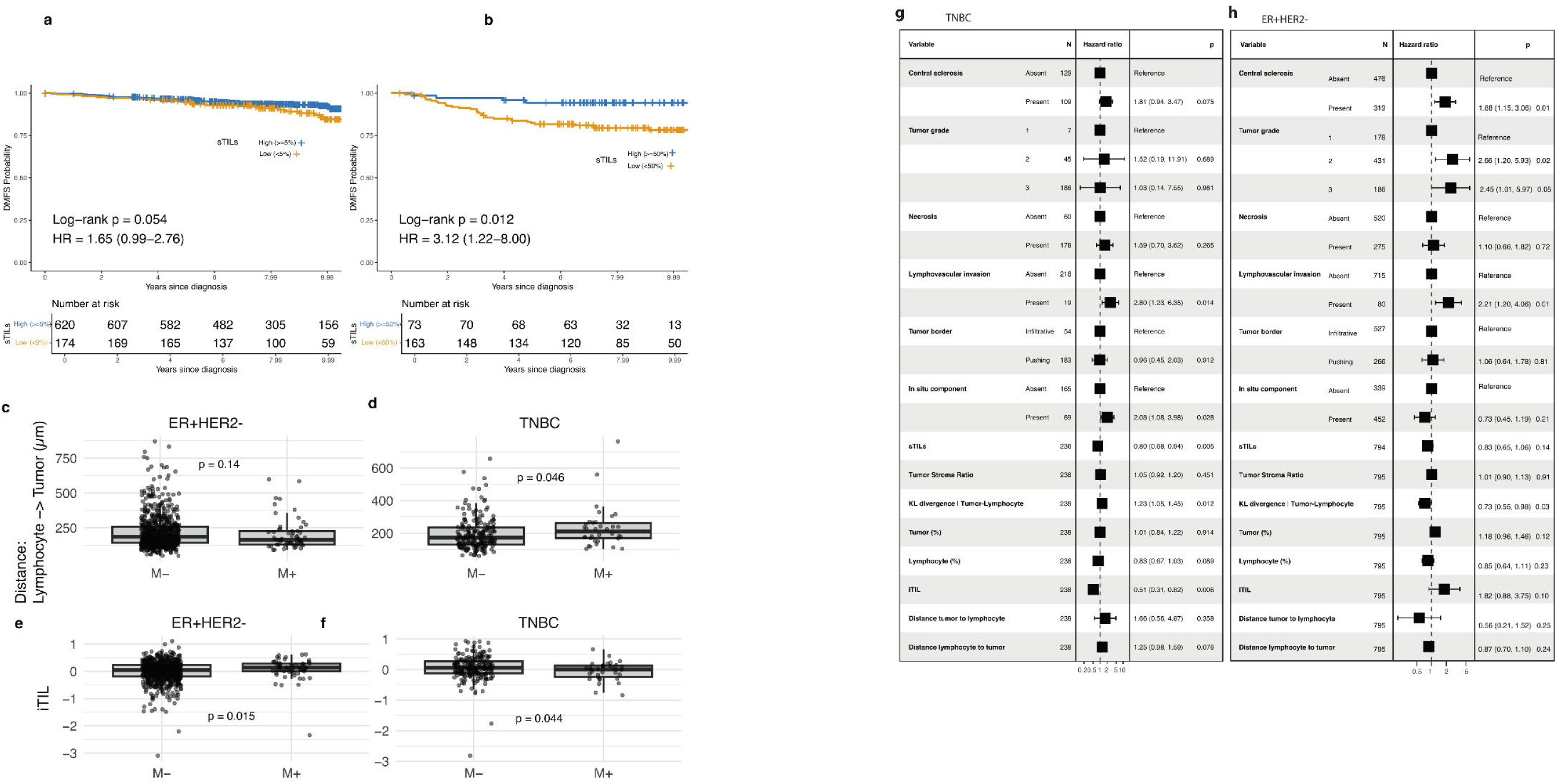
Prognostic modeling in breast cancer subtypes. a-b Distant metastasis–free survival (DMFS) by sTILs strata: ER⁺HER2⁻ >5% vs ≤5% (a), TNBC ≥50% vs <50% (b). Two-sided log-rank p values and Cox hazard ratios (HRs, 95% CIs) are annotated; x-axis in years. c-f; Tumor to lymphocyte distance (µm) by metastatic outcome for ER⁺HER2⁻ (c,e) and TNBC (d,f). Univariate cox models for variables in TNBC (g) and ER+HER2- (h). Box plots display median and IQR with whiskers at 1.5× IQR; Group comparisons use two-sided Wilcoxon rank-sum tests; survival comparisons in a–b use two-sided log-rank tests with HRs from Cox models. All tests are two-sided.

Model-derived lymphocyte densities were not prognostic in either subtype (**Supplementary Fig. 2d,e**). In TNBC, patients who developed metastases had greater lymphocyte→tumor distance (p = 0.046; **Fig. 2d**) and lower iTIL (p = 0.044; **Fig. 2f**), with the latter also prognostic in univariate analysis. In contrast, in ER⁺HER2⁻ tumors, iTIL was paradoxically higher in metastasis cases (p = 0.015; **Fig. 2e**). In line with this observation, an increase in KL divergence (less mixing) was adverse for TNBC patients, but beneficial in ER+HER2- patients (**Fig. 2g,h**). Among spatial and architectural features, lymphovascular invasion (LVI) was consistently adverse (ER⁺HER2⁻ HR 2.21, p = 0.01; TNBC HR 2.80, p = 0.01; **Fig. 2g,h**). Central sclerosis and a higher tumor grade was adverse in ER⁺HER2⁻ tumors, whereas the presence of an in situ component was adverse in TNBC (**Fig. 2g,h**). Neither necrosis nor an infiltrative versus pushing border showed prognostic impact in univariate analysis.

### Immune–architectural coupling by subtype and grade

We profiled how histologic architecture relates to immune abundance and spatial organization across TNBC, ER⁺/HER2⁻ grade 1–2, and ER⁺/HER2⁻ grade 3. Within each stratum, associations were ranked by absolute effect size with BH–FDR control (**Fig. 3a-b; Fig. S3a-d**). A conserved spatial–immune axis dominated: higher intratumoral lymphocyte infiltration (iTIL) tracked with greater tumor–lymphocyte mixing (lower KL) and shorter tumor↔lymphocyte distances, whereas longer distances aligned with less mixing and lower iTIL (**Fig. 3a–c**).

**Figure 3.**
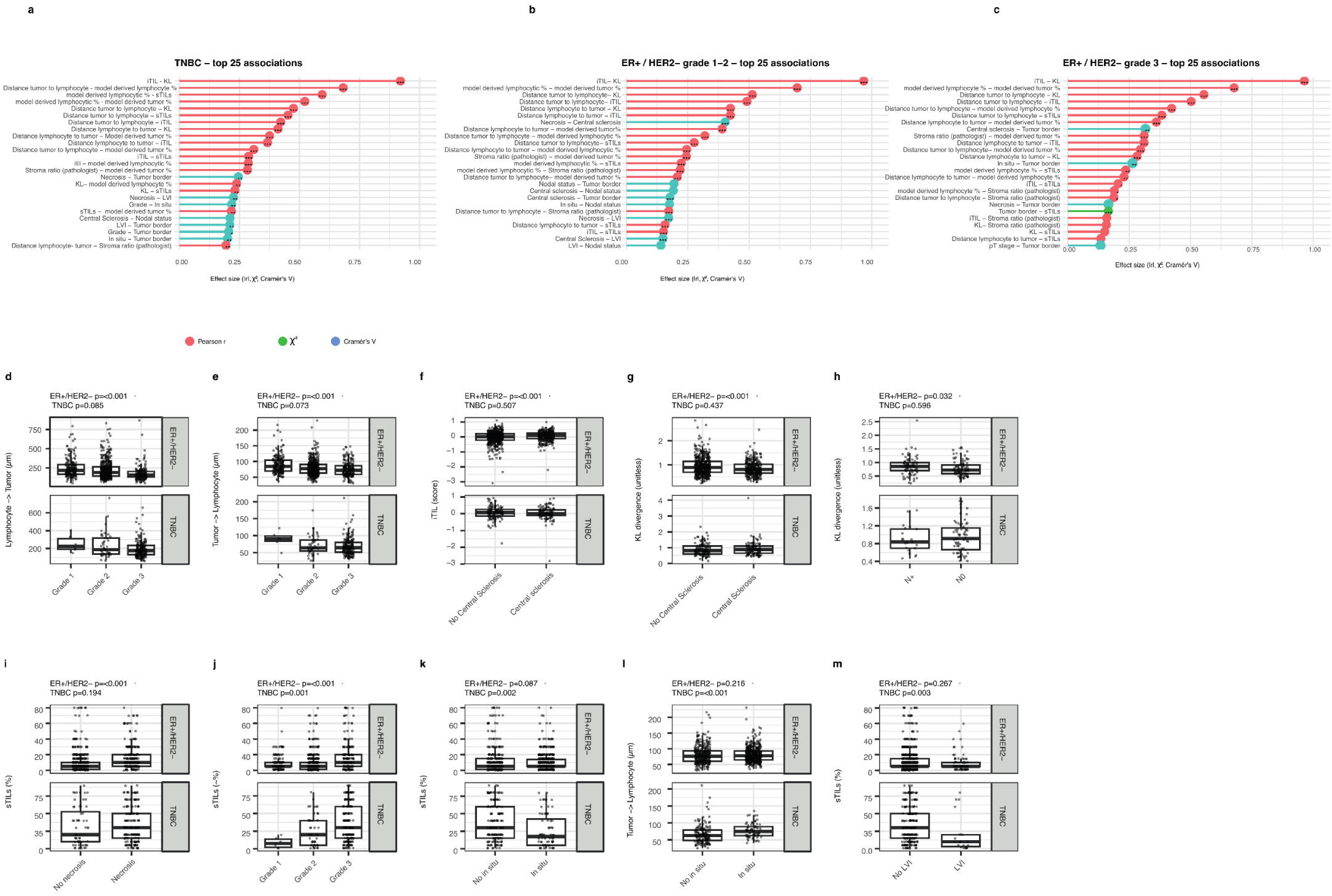
| Immune burden and spatial positioning across clinicopathologic features in ER+/HER2- and TNBC. a-c; Lollipop plots show the top 25 associations (by absolute effect size) within each subgroup: (a) TNBC, (b) ER+/HER2- grade 1–2, and (c) ER+/HER21 grade 3. Effect sizes are Pearson’s r for numeric–numeric pairs and η² for categorical-numeric contrasts. The x-axis gives the absolute magnitude of the association—|Pearson r| for numeric–numeric pairs (red), η² for categorical-numeric pairs (green), and Cramér’s V for categorical–categorical pairs (blue). Lollipop length encodes effect size; points are annotated with FDR-adjusted significance (“” q<0.001, “” q<0.01, “” q<0.05, “•” q<0.10; Benjamini–Hochberg). Self-pairs and mirrored duplicates were excluded. d-m; Boxplots show distributions of stromal TILs (sTIs %), intratumoral TIL score (iTIL; unitless), lymphocyte->tumor and tumor->lymphocyte nearest-neighbor distances (µm), and tumor–lymphocyte KL divergence (unitless) by feature category. Top row, ER+/HER2-; bottom row, TNBC. Medians are indicated by horizontal lines; points are individual cases (jittered). P-values are from Kruskal–Wallis tests per facet (panel); pairwise Wilcoxon tests are BH-adjusted. In ER+/HER2-, higher grade and stromal sclerosis associate with increased lymphocytic burden (sTILs/iTIL), altered positioning (both distances), and reduced separation (KL divergence). Node-negative cases show more inflamed, less segregated microenvironments. In TNBC, immune burden stratifies by grade, LVI, and in-situ component, with in-situ also showing larger tumor->lymphocyte distances, consistent with exclusion.

In ER⁺/HER2⁻ tumors, with increasing grade, lymphocyte–tumor distances shortened, sTILs increased, and KL decreased (grade-stratified contrasts FDR-significant; **Fig. 3d–j**; **Table S2-3**). Stromal sclerosis associated with higher iTIL and sTILs, a longer lymphocyte→tumor distance, and lower KL (all q<0.05; **Fig. 3d,e,j**). Node-negative tumors showed a more inflamed, less segregated milieu than node-positive (q<0.05; **Fig. 3h**). Necrosis aligned with higher immune burden and closer positioning (**Fig. 3i**). Because prognostic immune signals differed by grade (**Fig. S1d**), we re-tested architectural couplings in grade 1–2 and grade 3 separately (**Table S4**). In grade 1–2, architecture-driven signals were broad: both the grade step (2 vs 1) and sclerosis were significant across distances, iTIL, KL, and sTILs; node status showed smaller but consistent differences (KL, tumor→lymphocyte). In grade 3, the background appeared more uniformly inflamed; here lymphovascular invasion (LVI) became the dominant discriminator (iTIL, KL), and necrosis continued to stratify sTILs (q<0.001). In brief, architecture (grade, sclerosis) broadly modulates immune burden and geometry in ER G1–2, whereas in ER G3 the landscape is uniformly inflamed and LVI becomes the chief marker of reduced mixing and intratumoral infiltration; necrosis continues to mark higher stromal burden.

TNBC. sTILs increased with grade, while the in-situ component and LVI were associated with lower sTILs (q<0.01; **Fig. 3k,m**). In-situ disease also showed greater tumor→lymphocyte distance (q<0.001; **Fig. 3l**), consistent with an exclusionary phenotype.

In summary, ER⁺/HER2⁻ shows robust architecture-immune coupling—spanning grade, stromal sclerosis, and node status,while TNBC is primarily stratified by immune burden, with exclusion most evident when in-situ disease or lymphovascular invasion is present. Notably, the majority of architecture–immune associations were observed in ER⁺/HER2⁻ tumors (**Table S3**), underscoring the greater strength of coupling in this subtype.

### Multivariable modeling confirms subtype-specific drivers

We fit subtype-specific Cox models that jointly included clinicopathologic and immune features (**Table 3**). For TNBC (n=233; 37 events), after adjustment, sTILs remained independently protective (HR 0.82 per 10%; 95% CI 0.67–0.99; p=0.039). Tumor size (pT2–3 vs pT1) and LVI showed non-significant adverse trends, while grade, nodal status, in-situ component, iTIL, model-derived lymphocytic densities, and KL divergence were not independent predictors. In ER⁺/HER2⁻ (n=791; 65 events), grade (3 vs 1; HR 2.63, p=0.041; 2 vs 1; HR 2.34, p=0.040), LVI (HR 2.24, p=0.012), and central sclerosis (HR 2.09, p=0.0047) were independently adverse. KL divergence was protective (HR 0.43, p=0.026). iTIL showed a near-significant protective trend (HR 0.19, 95% CI 0.035–1.01; p=0.051), whereas sTILs, nodal status, tumor size, and model-derived lymphocytic densities were not significant in the multivariable setting. Taken together, immune burden (sTILs) captures the independent signal in TNBC, whereas in ER⁺/HER2⁻ disease architectural features (grade, sclerosis, LVI) and spatial context (KL) carry more prognostic weight.

**Table 3.** Multivariate Cox proportional hazards models evaluating associations between clinical, pathological, and immune-related variables and distant metastasis-free survival (DMFS) in patients with triple-negative breast cancer (TNBC; n = 233, 37 events) and estrogen receptor-positive/human epidermal growth factor receptor 2-negative (ER⁺HER2⁻; n = 791, 65 events) subtypes. Each subtype-specific model includes: tumor grade, nodal status (pN⁺ vs pN0), tumor size (pT2–3 vs pT1), lymphovascular invasion (LVI), stromal tumor-infiltrating lymphocytes (sTILs, per 10% increment), intratumoral TILs (iTIL), model derived lymphocytic densities, and KL divergence (a spatial separation metric). Central sclerosis was only included in the ER⁺HER2⁻ model, and in-situ component only in the TNBC model. Missing values were handled by listwise deletion. Hazard ratios (HRs) with 95% confidence intervals (CIs) are shown alongside Wald test p-values. Bold indicates statistical significance (p < 0.05). †p = 0.051 is near-significant.

| Variable | HR (TNBC) | 95% CI | p-value | HR (ER<sup>+</sup>HER2<sup>-</sup>) | 95% CI | p-value |
| --- | --- | --- | --- | --- | --- | --- |
| **Tumor grade 2** | 1.55 | 0.19–12.65 | 0.684 | 2.34 | 1.04–5.30 | 0.040 |
| **Tumor grade 3** | 1.52 | 0.20–11.65 | 0.686 | 2.63 | 1.04–6.63 | 0.041 |
| **Nodal status (pN<sup>+</sup>)** | 1.05 | 0.48–2.31 | 0.901 | 1.27 | 0.76–2.11 | 0.366 |
| **Tumor size (pT2–3 vs pT1)** | 1.79 | 0.93–3.48 | 0.083 | 1.30 | 0.76–2.22 | 0.343 |
| **Lymphovascular invasion (present)** | 2.18 | 0.87–5.47 | 0.098 | 2.24 | 1.20–4.19 | 0.012 |
| **Central sclerosis (present)** | — | — | — | 2.09 | 1.25–3.48 | 0.0047 |
| **In-situ component (present)** | 1.78 | 0.88–3.58 | 0.108 | — | — | — |
| **sTILs (per 10%)** | 0.82 | 0.67–0.99 | 0.039 | 0.86 | 0.66–1.13 | 0.279 |
| **iTIL** | 0.31 | 0.03–2.81 | 0.298 | 0.19 | 0.035–1.01 | 0.051† |
| **Model derived lymphocytic densities** | 1.07 | 0.80–1.43 | 0.650 | 0.93 | 0.79–1.09 | 0.345 |
| **KL divergence** | 0.84 | 0.40–1.74 | 0.631 | 0.43 | 0.205–0.905 | 0.026 |

Abbreviations: HR, hazard ratio; CI, confidence interval; TNBC, triple-negative breast cancer; ER, estrogen receptor; HER2, human epidermal growth factor receptor 2; LVI, lymphovascular invasion; sTILs, stromal tumor-infiltrating lymphocytes; iTIL, intratumoral TIL score (negative product of model derived lymphocytic densities and normalized tumor–lymphocyte KL divergence); KL, Kullback–Leibler divergence.

### Multiplex Phenotyping Reveals Functional and Spatial Constraints on Immune Control in TNBC

To test whether H&E-derived immune quantity and spatial metrics reflect functional cell states and microanatomic constraints, we profiled a case–control matched TNBC subset (n=99; 33 metastatic; **Fig. 4**) by multiplex immunodetection (**Fig. 4a-b)** and linked these data to H&E features (**Fig. 4 a,n**) and bulk RNA sequencing (**Fig. 4d**). Transcriptomic analysis showed downregulation of inflammation-related signatures (IFN-γ, IFN-α, TIS, and lymphocytic signatures) and reduced tissue-resident memory T-cell (Trm)^37^ signals in patients who developed metastases. Conversely, hedgehog signaling, TGF-β, and stromal programs were enriched in these patients.

**Figure 4.**
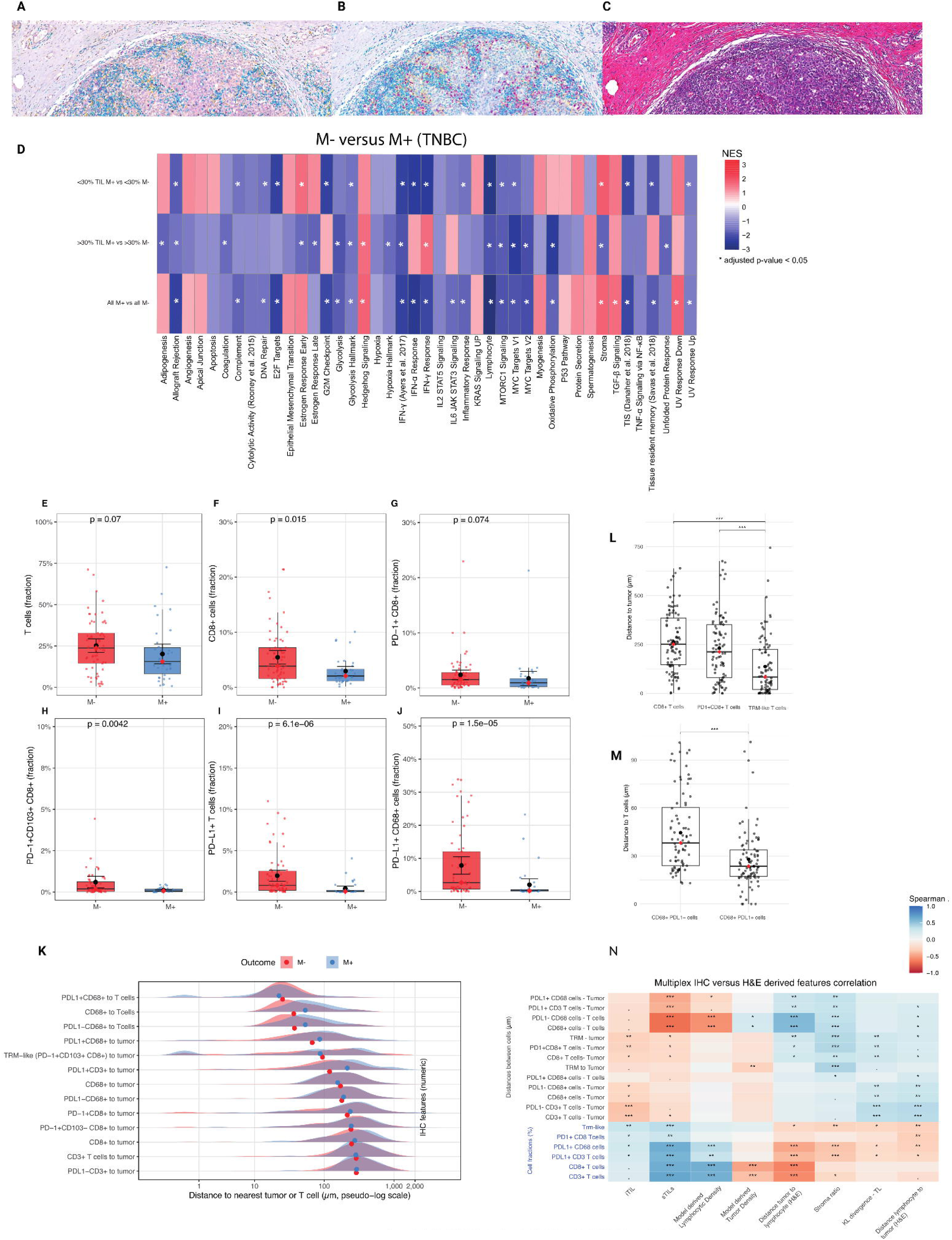
| Multiplex immunophenotyping in TNBC. a–c, Representative multiplex IHC images: PD-1 (yellow), CD8 (teal), CD103 (purple) (a); PD-L1 (yellow), CD3 (teal), CD68 (purple) (b); corresponding H&E (c). d, Gene set enrichment analysis (GSEA) of bulk RNA sequencing data from 63 TNBC patients, focusing on cancer hallmarks and selected gene signatures. Normalized enrichment score (NES) is shown for each comparison and signature. Significantly enriched signatures are marked with an asterisk. e–j, Fractions of immune subsets in TNBC patients with (M⁺) or without (M⁻) metastasis (n=97): CD3⁺ T cells, CD8⁺ T cells, PD-1⁺CD8⁺ T cells, CD103⁺PD-1⁺CD8⁺ TRM-like cells, PD-L1⁺ CD3⁺ T cells and PD-L1⁺CD68⁺ myeloid cells. k, Distribution of multiplex IHC–detected cells by distance to tumor or T cells. l, Distances to tumor for CD8⁺ vs CD8⁺PD-1⁺CD103⁺ TRM-like cells. m, Distances to T cells for CD68⁺ vs PD-L1⁺CD68⁺ myeloid cells. n, Spearman correlations between IHC derived features and H&E derived features on whole slide analyses. Boxplots show minimum, maximum, median, and IQR, with medians marked by red dots. Comparisons in e–j used two-sided Wilcoxon rank-sum tests.

These differences persisted even among tumors with <30% sTILs, indicating that microenvironmental alterations extend beyond overall lymphocyte burden (**Fig. 4d**). We therefore assessed markers of immune activation and checkpoint regulation in whole-slide images: CD103, PD-1, and CD8 to identify Trm-like cells^38,39^, and PD-L1, CD68, and CD3 to evaluate immune checkpoint presence, given that PD-L1 in breast cancer is primarily expressed on immune rather than tumor cells ^40^ (Fig. 4a–c; Fig. S4). Total CD3⁺ T-cell density did not differ by outcome (p=0.07, **Fig. 4e**). In contrast, cytotoxic and Trm-like subsets were lower in metastatic patients, including CD8⁺, PD-1⁺CD8⁺, and CD103⁺PD-1⁺CD8⁺ tissue-resident memory–like cells (**Fig. 4g–i**). Spatial analysis showed that PD-1⁺ and CD103⁺ T cells localized closer to tumor cells than total CD3⁺ or CD8⁺, with PD-1⁺CD103⁺CD8⁺ cells in closest proximity (p = 9.4 × 10⁻⁶ vs PD-1⁺CD8⁺; p = 5.2 × 10⁻¹⁰ vs CD8⁺; Fig. 4l). Intercellular distances did not differ by outcome, suggesting that numerical loss rather than relocalization underlies metastatic progression. Focusing on PD-L1 expression, PD-L1⁺ CD3⁺ T cells were significantly reduced in patients who developed metastases (Fig. 4f), and PD-L1 expression on CD68⁺ macrophages was likewise lower in metastatic patients (Fig. 4j). Notably, PD-L1⁺CD68⁺ cells localized closer to T cells than PD-L1⁻CD68⁺ cells (Fig. 4m), consistent with localized immunomodulation.

H&E-derived metrics recapitulated multiplex IHC geometry (Fig. 4n). Higher sTILs correlated with greater T-cell and CD8⁺ density and with shorter T-cell→tumor distances (q ≤ 0.001). Slides with higher sTILs also showed PD-L1⁺ T cells and PD-L1⁺CD68⁺ macrophages positioned closer to tumor (q ≤ 0.001). H&E spatial features reflected these patterns: higher KL divergence aligned with longer T-cell→tumor distances (q ≤ 0.01), whereas higher iTIL associated with shorter distances (q ≤ 0.05).

Stromal architecture exerted additional constraints. A higher stroma ratio correlated with greater tumor→PD-1⁺CD8⁺ and CD68⁺→T-cell distances (q ≤ 0.01). CD68⁺→T-cell spacing increased in parallel with H&E tumor→lymphocyte separation, suggesting that lymphoid and myeloid positioning are governed by shared architectural barriers. Conversely, TRM-like CD8⁺PD-1⁺CD103⁺ cells correlated with shorter tumor→lymphocyte distances and lower stroma ratio. Together, these findings indicate that stromal abundance on H&E is linked to altered immune–tumor geometry, reflected in both lymphoid and myeloid compartments.

### High-dimensional spatial proteomics of immune-dense foci in TNBC

To complement H&E-derived metrics, we profiled pathologist-selected, immune-tumor dense regions from TNBC using imaging mass cytometry (IMC; 1.5-mm cores; n = 89; outcome-blinded; **Fig. 5a**, **Supplementary Fig. 5a**). Our TMA sampling strategy was designed to enrich tumor–immune interaction zones rather than provide a proportional overview of tumor-wide infiltration. This increases sensitivity for studying tumor–immune intermixing but precludes interpretation as a measure of overall immune burden.

**Fig. 5.**
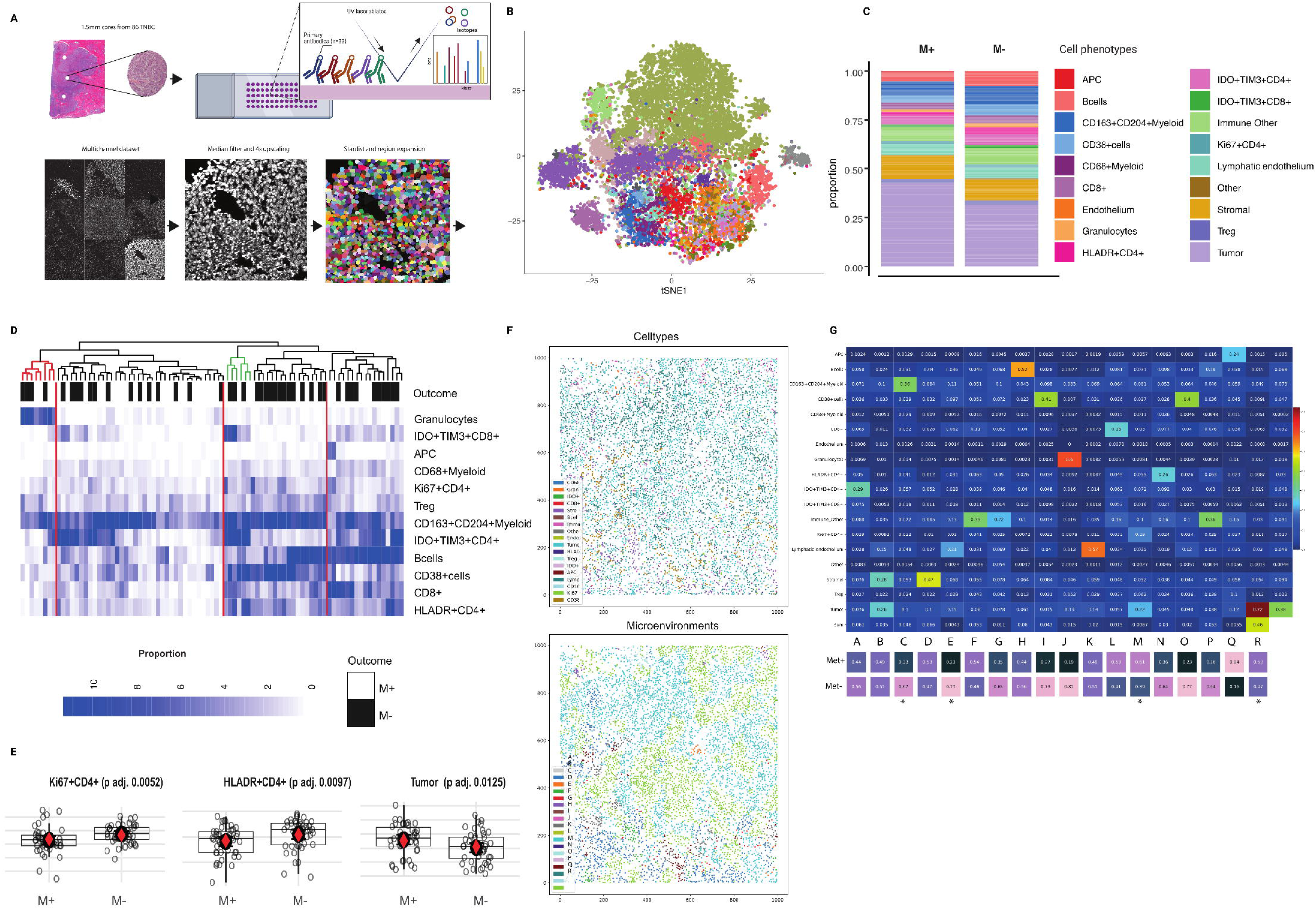
| Immune-tumor dense areas in TNBC patients with and without subsequent distant metastasis. a) Construction of TMA with 1.5mm cores preselected as having representative immune infiltration in immune dense areas by a dedicated pathologist. The DNA channel was used for segmentation. Images were denoised and 4x upscaled. StarDist nuclei recognition and region expansion was used, resulting in ∼1 million cells segmented. b) TSNE map from the 18 cell clusters. Each cluster has a unique color assigned (bar on the right). Values in the brackets next to the cluster numbers indicate the relative size of clusters. c) Relative proportions of all cell phenotypes in patients with (M+) and without (M-) metastatic progression. d) Heatmap of cell levels across all TNBC patients, M+ and M- status annotated on top with a dendrogram from hierarchical clustering. e) Comparison of cell type proportions across M+ and M- patients. The logit transformation (logit(p)=log(p/1−p)) was applied before performing the Wilcoxon rank-sum test to transform the cell proportions to a scale that can be analyzed using methods that assume normality. f) Example of cell phenotype map and microenvironmental map of one sample. g) Microenvironments (MEs) retrieved from spatial-LDA analysis. The five MEs on the right side, annotated with *were statistically different between M+ and M- TNBC patients (Kruskal-Wallis). Boxplots in e display minimum (Q0), maximum (Q4), median (Q2), and interquartile range, with the median indicated by a red dot.

A 21-marker panel^41^ with single-cell segmentation^42^ identified 18 expected immune and stromal phenotypes (**Fig. 5b**, **Supplementary Fig. 5**). Unsupervised clustering of core-level compositions yielded four groups without separation by metastatic outcome (**Fig. 5d**), suggesting no gross compositional shift in immune-dense areas. One cluster consisted of tumor–immune mixing zones with granulocyte infiltration, another was enriched for IDO⁺TIM3⁺ CD4⁺ T cells, a third contained predominantly CD163⁺CD204⁺ myeloid cells, and a fourth showed CD163⁺CD204⁺ myeloid cells together with CD38⁺ B cells. Despite similar overall compositions, patients who developed metastases had greater tumor-cell content in these regions and fewer Ki67⁺ and HLA-DR⁺ CD4⁺ T cells (**Fig. 5e**).

To interrogate microanatomic patterning, we applied spatial-LDA ^43^ and derived 18 microenvironment elements (MEs) from cell-type co-occurrence (**Fig. 5g**). Four MEs differed by outcome. Progressors were enriched for a tumor-rich ME (R) and an ME of tumor cells admixed with proliferating CD4⁺ T cells (M), despite an overall reduction of proliferating CD4⁺ cells at the ROI level (**Fig. 5d–e**). Non-progressors were enriched for ME-C, defined by CD163⁺CD204⁺ myeloid cells, and ME-E, characterized by lymphatic endothelium (D2-40).

Together, these results in this relatively small pilot indicate that outcome differences in TNBC are not reflected by bulk immune counts within immune-dense foci but instead by specific tumor-centric microenvironments. In other words, prognosis is shaped less by “how much immune infiltrate is present” and more by “where and how immune subsets are organized.”

## Discussion

This multicenter study of young women with early breast cancer integrates expert pathology, deep learning on whole-slide H&E images, and multiplex immunophenotyping to dissect how immune quantity and spatial organization relate to distant metastasis-free survival (DMFS). Prognostic immune signals are subtype specific: in triple-negative breast cancer (TNBC), immune burden captured by stromal tumor-infiltrating lymphocytes (sTILs) dominates, whereas in ER⁺/HER2⁻ disease, tissue architecture and spatial separation contribute more independent information than bulk immune density.

Consistent with prior literature^2,44,45^, higher sTILs were strongly associated with improved DMFS in TNBC, including after adjustment for clinicopathologic covariates and in chemotherapy-treated patients. In fully adjusted models, sTILs were the only independent immune feature, indicating that in immunologically active TNBC, lymphocytic infiltration captures the principal prognostic signal. In ER⁺/HER2⁻ tumors, by contrast, sTILs were protective mainly in grade 1–2 and lost independence once architectural covariates were included, suggesting that in luminal cancers, measured immune infiltration may in part reflect tumor-intrinsic biology (for example, proliferation or differentiation state) rather than a dedicated antitumor response.

A strength of our study is standardized single-cell spatial profiling directly from routine H&E slides. Our cohort-trained pipeline produced harmonized readouts: cell fractions/densities, bidirectional nearest-neighbor distances, and Kullback-Leibler (KL)^46^ divergence as a measure of immune–tumor separation, from which we defined iTIL, a mixing-aware metric coupling lymphocyte abundance with colocalization. In TNBC, lower separation (lower KL with greater colocalization) and higher iTIL aligned with better outcomes, consistent with an inflamed, interdigitated immune niche. In ER⁺/HER2⁻ tumors, higher KL divergence was independently protective, implying that in low-inflammation contexts, spatial separation may reflect architectural containment rather than immune exclusion. These patterns argue against one-size-fits-all interpretation of spatial metrics across breast cancer subtypes.

Model-derived lymphocyte density correlated with sTILs, particularly in TNBC, but was less prognostic than pathologist scoring. Expert review implicitly encodes spatial context ^4^, including weighting of the tumor-stroma interface, discounting of necrotic or in situ regions, and integration of border morphology, which model derived cell densities misses. Concordance was stronger in TNBC, where abundance and mixing align, and weaker in ER⁺/HER2⁻, where architecture modulates immune positioning. Notably, some spatial patterns previously described with CD8⁺ staining^4,8,9^ were rarely identifiable on H&E by pathologists, which does not distinguish CD8⁺ from other lymphocyte subsets. This highlights the need for region-aware computational models that explicitly account for epithelial–stromal boundaries and other biologically relevant compartments.

In TNBC, immune features are tightly correlated with one another and only weakly with classic morphologic descriptors, suggesting that immune burden subsumes most prognostic information. In ER⁺/HER2⁻, particularly grade 3, border configuration and central sclerosis tracked with both immune quantity and separation, underscoring that microanatomy shapes immune organization. Multivariable models highlighted clinical weight: grade, lymphovascular invasion (LVI), and central sclerosis were independently adverse, while spatial separation added signal beyond bulk density. These results support integrated biomarkers that combine architecture with immune metrics in luminal cancers.

Multiplex immunophenotyping added mechanistic depth. Total CD3⁺ did not differ by outcome, but effector-skewed, tumor-reactive subsets (CD8⁺, PD-1⁺CD8⁺, and CD103⁺PD-1⁺CD8⁺ TRM-like cells) were lower in patients who metastasized. These subsets localized closest to tumor cells across the cohort, and their distances did not vary by outcome, indicating that reduced abundance, rather than relocalization, distinguishes poor-outcome tumors. PD-L1⁺CD68⁺ myeloid cells, which sat nearer T cells than PD-L1⁻ counterparts, were likewise reduced in metastasizing cases, consistent with reprogrammed immune and myeloid crosstalk in aggressive disease. H&E-derived spatial metrics mirrored these geometries: higher KL aligned with longer T-cell to tumor distances on IHC, while higher iTIL tracked with closer apposition, and stromal content modulated myeloid–lymphoid spacing. These cross-modal concordances confirm that H&E spatial readouts capture real microanatomic constraints experienced by functional immune subsets.

We did not perform a dedicated inter-observer reproducibility analysis; however, all histopathological features were centrally scored by six breast pathologists using validated frameworks with demonstrated reproducibility in prior studies. The cohort is observational and restricted to women ≤50 years, which limits generalizability, and reflects 2004 to 2011 treatment (pre-neoadjuvant chemotherapy and immunotherapy). TNBC event counts were modest; null findings for some spatial metrics should be interpreted cautiously. The deep-learning pipeline may be sensitive to training distributions and region definitions, which necessitates external validation. Interactions with genomic assays used in ER⁺/HER2⁻ disease (for example, MammaPrint ^47^) were not assessed. Cancer-associated fibroblasts (CAFs) were not characterized; prior work implicates CAF subsets in immune exclusion, therapy response and prognosis ^19,20,48,49^, but fibroblasts are morphologically heterogeneous and not reliably identifiable on H&E. Tissue microarray sampling intentionally enriched tumor–immune interaction zones rather than providing proportional mapping of all regions, which may underrepresent immune-poor areas; complementary whole-slide H&E analyses were therefore used to assess broader immune distribution. Finally, H&E spatial patterns are proxies and cannot resolve e.g. CD8⁺ specifically; low-plex IHC partly addresses this but remains exploratory.

Two lines of work are warranted. First, independent validation of architecture-aware spatial metrics (iTIL and KL divergence) and outcomes in ER⁺/HER2⁻ disease is needed, using prespecified thresholds and harmonized pipelines. Second, these features should be tested in contemporary cohorts where the tumor–immune milieu is perturbed (neoadjuvant chemotherapy, immunotherapy, or combinations) to evaluate persistence of prognostic signal and potential predictive value. Methodologically, H&E analytics should be refined to infer functional immune states (for example, antigen-presenting niches and tumor-reactive T cells) and to quantify architectural fitness, anchored to minimal bright-field panels and calibrated against multiplex IHC or spatial transcriptomics, particularly to resolve fibroblast subsets and stromal constraints on immune access.

Immune metrics are best interpreted through subtype and architectural context. In TNBC, immune burden and tumor–immune colocalization carry the strongest prognostic signal; in ER⁺/HER2⁻ disease, architectural context and spatial separation are more informative. Standardized spatial features from routine H&E, complemented by targeted multiplex phenotyping, provide a feasible basis for integrated biomarkers that can refine risk stratification and, pending validation, inform therapy selection in early breast cancer.

## Methods

### Patient Cohort and Study Design

Patients were drawn from the Young Boost Trial (YBT; NCT00212121)^27,28^, a multicenter randomized study conducted in the Netherlands, France, and Germany (2004–2011). Eligible participants were ≤50 years at diagnosis with primary invasive breast cancer without distant metastasis (M0). Neoadjuvant systemic therapy was not permitted, ensuring all surgical material was treatment-naïve. Clinical follow-up (locoregional recurrence, distant metastasis, overall survival) was available through February 22, 2019.

Formalin-fixed paraffin-embedded (FFPE) primary tumor blocks were requested from all participants. Of 1,481 specimens retrieved, 1,370 were evaluable. Distant metastasis-free survival (DMFS) was defined as the interval from randomization to metastasis or death, with censoring at last follow-up. Among evaluable tumors, 240 were triple-negative breast cancers (TNBC; ER and PR <10% and HER2– by national guidelines), and 797 were ER⁺HER2⁻.

### Ethical Approval

The study followed Good Clinical Practice and the Declaration of Helsinki. The protocol and informed consent were approved by institutional review boards. All patients gave written informed consent. Translational analyses were approved by the Netherlands Cancer Institute IRB (CFMPB27).

### Histopathological Review

Whole-slide H&E sections (n = 1,370) were centrally reviewed by six breast pathologists using SlideScore^30^. The following features were assessed: histological subtype and grade (Bloom–Richardson^50^); tumor border; central sclerosis/fibrosis; necrosis; tumor–stroma ratio; immune hotspots; immune-excluded phenotype; and stromal tumor-infiltrating lymphocytes (sTILs). ER, PR, and HER2 status were determined by immunohistochemistry; HER2 FISH/SISH results superseded IHC when available. Triple-negative breast cancer (TNBC) was defined as ER and PR <10% and HER2 negative.

**Tumor border** was classified as infiltrative or pushing, based on the proportion of the tumor circumference with a pushing growth pattern, consistent with prior studies linking border morphology to prognosis in invasive breast cancer^33,51–54^.

**Central necrosis/fibrosis** was recorded when necrotic or fibrotic tissue was present in the tumor center. Central fibrosis has been associated with stromal remodeling and immune exclusion ^13,32,54,55^.

**Tumor necrosis** was evaluated across the entire tumor area. Presence of necrosis has been consistently associated with aggressive biology and adverse outcomes. ^56^

**Tumor–stroma ratio (TSR)** was visually estimated as the percentage of stromal tissue within the tumor bed and categorized as stroma-poor (<50%) or stroma-rich (≥50%). TSR is a validated prognostic marker across multiple solid tumors^57,58^.

**Immune hotspots** were defined as clusters of densely packed lymphocytes identifiable on H&E sections. These regions may represent areas of localized immune activity, including immature tertiary lymphoid structures (TLS); however, definitive identification requires immunohistochemistry. ^59^

**Immune-excluded phenotype** was defined as lymphocytes restricted to the peritumoral stroma. As this pattern was observed at very low frequency in our cohort, it was not included in subsequent analyses. We note that immune exclusion, as previously described ^8,9^, has primarily been characterized using CD8 immunohistochemistry rather than H&E.

**sTILs** were quantified as the percentage of stromal area infiltrated by mononuclear immune cells (0–100%), following the international guidelines ^5^.

### H&E Automated Analysis: Cell Type Classification

Our automated analysis of H&E slides targeted lymphocytes and tumor cells through a two-step process: locating cell nuclei and classifying their types (**Fig. 1a**). For the first task, we used a U-Net architecture with a pretrained ResNet 152 backbone for locating the cell nuclei positions over the entire WSI tissue. Once done with locating the cells, a DenseNet 161 architecture was selected for the cell type classification model. Training of these models required two separate datasets: one with densely annotated nuclei positions for the first task and another sparsely annotated with cell type labels for the second task. Both datasets were annotated and labeled by expert pathologists. The advantage of the two step approach for cell H&E cell classification is that it allows fine-tuning the classification model with minimal new labeled cells, since the exhaustively annotated nuclei locations are no longer required for the classification training. This saves the time required for augmenting the classification training data by the pathologist allowing to extend the data to more slides and tissue samples and increasing the generalization power.

The locating model was trained on 256x256 patches with overlapping margins (32 pixels) to ensure comprehensive cell nuclei detection without double-counting of the boundary cells. Training was done with the help of a simple distillation process ^60^ using StartDist ^61^, a pre-trained cell segmentation model on H&E as the teacher. StarDist outputs were smoothed with region growing and augmented with additional nuclei position annotations from pathologists that were grown to the size of a small cell to include as many as possible (**Fig. 1b**). The training loss function combined terms for these grown labels and teacher softmax output. Cell locating performance was evaluated heuristically by measuring distances on an unseen annotated test set and achieved an accuracy of over 98.6%.

Each detected cell nuclei location was then extracted as the center of a 256x256 sample at 0.5 micron per pixel zoom level, for training (and inference) of the classification model (**Fig. 1c**). By providing a bigger context for identifying the cell types, the extra margin of tissue helped improve our performance in our initial tests, compared to smaller margins. To ensure generalizability of our results, our models were only trained on a selection of slides from the ICGC ^62^ cohort, and then applied to score the target YoungBoost cohort. The target cohort was completely unseen in the training process. Prioritizing classification of tumor cells, lymphocytes, and normal tissue (including epithelial cells), our model achieved an overall classification accuracy of 97%. Other cell types such as other immune classes, red blood cells, fibroblasts, fat cells and nerve cells were all regarded as normal (**Fig. S4a**). The overall F1-score of the 3-way classification model was 0.983 calculated by using what is called the micro averaging strategy, which means that precision and recall were based on the sum of total TP, FP, and FN for all 3 classes. In contrast to prior CD8-based spatial phenotyping approaches^8,9^, our H&E-derived spatial patterns serve only as proxies of immune localization and cannot specifically distinguish e.g., CD8⁺ T cells from other lymphocyte subsets.

### Multiplex Immunohistochemistry Immunodetection and Analysis

The antibody panels for multiplex IHC were designed to address two complementary axes: (i) cytotoxic T-cell activation and exhaustion (CD8, CD103, PD-1) and (ii) the interface between T cells, myeloid cells, and immune checkpoints (CD3, CD68, PD-L1). In addition, we aimed to construct smaller panels that would allow spatial information to be captured while remaining compatible with bright-field microscopy and scalable to larger cohorts. Although exploratory, these panels were guided by prior evidence implicating CD103⁺CD8⁺ T cells^23,24,63^ in tissue-resident immunity and APC/PD-L1 interactions in immune evasion^40,64,65^.

We performed multiplex immunohistochemistry on 99 TNBC patient samples (all E-cadherin+), using markers CD103/PD-1/CD8 and PD-L1/CD68/CD3 (**Fig. S2d**). Immunodetection was conducted using a Discovery Ultra automated stainer, following standard protocols for paraffin sections. For For the PD-1 (Yellow), CD8 (Teal) and CD103 (Purple) panel, PD-1 was detected first using clone NAT105 (RTU, 16min RT, Roche), followed by CD8 detection with clone C8/144B (1/200, 32 min 37C, Agilent/DAKO), and CD103 with clone EPR4166(2) (1/500 dilution, 1 hour at 37C, AbCam). For the PD-L1 (Yellow), CD3 (Teal) and CD68 (Purple) panel, PD-L1 was detected first using clone 22C3(1/40 dilution, 1 h 37C, Agilent/DAKO), followed by CD3 detection with clone SP7(1/100, 32 min 37C, ThermoScientific), and CD68 with clone KP1 (1/10000, 32 min 37C, Agilent/DAKO). Each antibody was visualized using appropriate secondary antibodies and detection kits from Ventana Medical Systems. Slides were counterstained with hematoxylin.

The slides were digitized using the PhenoImager HT system (Akoya Bio). RGB overview scans at 10x magnification (1.0 um/pixel) were used to select regions of interest, which were then scanned at 20x magnification (0.5um/pixel) using MSI technology from Akoya. Each 20x field is imaged using a Liquid Crystal Tunable Filter (LCTF) to capture the whole visible spectrum for each pixel. The spectral library was created using slides stained with single colors. The slides were processed with inForm (v2.5.1, Akoya Bio) to separate each color into a separate channel. The resulting multi-layered TIFF files were loaded into HALO (v3.2, Indica Labs) and fused to generate whole slide images.

Digitized slides were analyzed using HALO software, with specific convolutional neural networks (CNNs) trained for cell detection and classification. The pipeline ran in two threads: the first model segmented cell nuclei and the second classified cell types. The tumor bed was manually annotated to exclude immune cells detected outside this region, ensuring accurate distance measurements. Pathologists annotated the tumor bed in https://SlideScore.com ^66^, and detections outside the tumor bed were excluded using QuPath^67^. This resulted in 103.019.815 cells. We measured mean nearest neighbor distances (mNND) between various cell types using Python (v3.9.12, scikit-learn v1.0.2, pandas v1.4.2). For the CD3/CD68/PD-L1 dataset, mNNDs were measured between CD3^+^ to tumor cells, CD3^+^PD-L1^+^ to tumor cells, CD3^+^PD-L1- to tumor cells, CD68^+^ to tumor cells, CD68^+^PD-L1^+^ to tumor cells, CD68^+^PD-L1- to tumor cells, CD68^+^ to T cells, CD68^+^PD-L1^+^ to T cells, and CD68^+^PD-L1^-^ to T cells. For the CD8/CD103/PD-1 dataset, mNNDs were measured between CD8^+^ to tumor cells, CD8+PD-1+ to tumor cells, CD8^+^PD-1^-^ to tumor cells, CD8^+^PD-1^+^CD103^+^ to tumor cells, and CD8^+^PD-1^+^CD103- to tumor cells.

### Imaging Mass Cytometry

For primary resections, up to three distinct 1.5-mm cores were obtained per patient. Core selection was guided by paired H&E slides centrally reviewed by a breast pathologist, with the aim of capturing representative tumor morphology (growth pattern, histology, and grade) as well as regions enriched for lymphocytic infiltration when present. In cases of uniform infiltration, tumor-focused cores inherently reflected the immune contexture. Regions composed predominantly of fat, fibrosis, necrosis, or in situ carcinoma were excluded. For biopsies, up to three cores were sampled using the same criteria. This strategy enriched tumor–immune interaction zones while minimizing non-informative tissue.

Antibodies were conjugated using the MaxPar antibody labeling kit and protocol (Fluidigm). Following conjugation, antibodies were eluted in 50 μl W-buffer (Fluidigm) and stabilized with 50µl antibody stabilizer (Candor Bioscience, Wangen im Allgäu, Germany). The antibody panel is based on previously published work ^41^ and targets epitopes specific for the tumor microenvironment. Tissue samples were formalin- fixed and paraffin embedded at the participating center of surgery for the Young Boost Trial. Tissue cores with a diameter of 1,500 μm were punched from three tumor regions containing both tumor cells and immune infiltrations. Regions of interest were selected by a trained pathologist based on hematoxylin and eosin stains performed on consecutive tissue sections. Areas of 1,000 × 1,000 μm were then ablated and acquired at 200 Hz using the Hyperion mass cytometry system. For each patient, two cores were analyzed. Data from four tissue microarray (TMA) images were exported as MCD files and visualized using the Fluidigm MCDTM viewer. A total of 86 ROIs were included in the downstream analysis. To better distinguish antibody signal from noise, each marker was visually inspected, and minimum signal thresholds ranging from 1 to 10 dual counts were set using the Fluidigm MCDTM viewer. Raw data were exported as OME-TIFF files, and thresholds were saved as plain text files.

Cells were segmented using the StarDist model within Fiji ^68^. The pretrained “Versatile (fluorescent nuclei)” model of StarDist ^61,69^ was used to segment cells, requiring a nuclear signal. The IMC dataset contained two DNA channels labeled with Ir(191) and Ir(193). To reduce noise, both channels were summed, and a median filter with a radius of 0.5 px was applied. The original pixel size of 1 μm/pixel was too large for StarDist and required upscaling 4x to obtain a pixel size of 250 nm/pixel. Subsequent 4x downscaling of the segmentation results produced a label map containing a region for each nucleus. Using the CLIJ package in Fiji ^70^, which facilitates parallel processing on the GPU, regions were expanded by 2 pixels to include the cytosol. Intensities were normalized per image and channel according to the threshold file. The mean intensity for each region was measured, and results were exported as CSV files. This workflow integrates classical image analysis and deep learning and is freely available online.

Cell clustering was performed using the FlowSOM ^71^ and ConsensusClusterPlus^72^ packages, following the CyTOF workflow described by Nowicka et al. ^42^. Single-area CSV files were loaded into an FCS flowset, and data were arcsinh transformed using the flowCore package with a cofactor of 5. An expression matrix from each element of the flowSet object was created to enable analysis of marker expression across all samples. Twenty lineage markers and twelve functional markers were used for downstream analyses. A self-organizing map (SOM) was constructed, assigning cells to 100 grid points based on similarity. Meta-clustering and expert-based merging of clusters, followed by re-clustering of T cell and myeloid clusters, resulted in the final cell type assignment shown in 2-dimentional t-SNE mapping (**Fig. 5b**).

The cell type assignments were laid out on the TMA samples. Each TMA and the annotated cell locations were processed as a document to fit a spatial-LDA ^43^ model. The model allocated the cells to 18 tumor microenvironments with different compositions of cell types (**Fig. 5g**) based on cell type population and proximity (**Fig. 5f**). The significance of the microenvironment’s expression difference between the M+ and M- groups was then tested using the Kruskal-Wallis test finding significance cases in four of the 18 MEs.

### RNA seq analyses

Reads were mapped to the transcriptome using Tophat ^73^. Subsequently, gene expression was determined by counting the reads that were mapped uniquely to the transcriptome using Icount, based on HTSeq-Count ^74^. Lowly expressed genes with a total count across all samples less or equal to 10 were filtered out. Further analysis was performed using DESeq2. Data were transformed using a variance stabilizing transformation (vst), and further exploration was done using a principal component analysis. Differential gene expression analysis was done using the DESeq function from the DESeq2 package using a contrast between M^+^ and M^-^ patients. Hierarchical clustering was done using the euclidean distance between samples at the gene and group level, taking the top 100 differentially expressed genes (DEGs). For the gene set enrichment analysis (GSEA), we used gseaMultilevel function from the R Bioconductor package “fgsea” package and the statistics provided by the results from DESeq2 to rank genes. We preselected gene signatures of interest ^24,75–77^ and significant signatures were selected using a threshold of adjusted p-value of 0.05.

### Statistical analyses

Group comparisons were performed using Kruskal–Wallis or Wilcoxon rank-sum tests for continuous variables and χ² tests for categorical variables. Survival outcomes were assessed with Kaplan–Meier estimates and log-rank tests. Univariable and multivariable Cox proportional hazards models were used to evaluate prognostic factors. Because proportional hazards assumptions were violated in pooled analyses, all models were fit separately for TNBC and ER⁺HER2⁻ subtypes.

Distant metastasis–free survival (DMFS) was defined as time from diagnosis to distant recurrence or death, with censoring at last follow-up. For univariable analyses, prespecified clinicopathologic and spatial features were tested (tumor grade, nodal status, tumor size, lymphovascular invasion [LVI], central sclerosis or in-situ component, sTILs, iTIL, model derived lymphocytic densities, KL divergence, and distance metrics). Hazard ratios (HRs) with 95% CIs are reported; sTILs were modeled per 10% increase.

We fitted subtype-specific Cox proportional hazards models for distant metastasis–free survival (DMFS) in triple-negative breast cancer (TNBC; n=233, 37 events) and ER⁺/HER2⁻ (n=791, 65 events). DMFS was analyzed as a time-to-event endpoint using the definitions provided in the study dataset; individuals without a DMFS event were censored at last follow-up.

Covariate inclusion did not differ by univariable significance; all pre-specified variables were entered jointly. Missing values were handled by listwise deletion within each subtype model (i.e., complete-case analysis). Model coefficients are reported as hazard ratios (HRs) with 95% confidence intervals (CIs) and Wald test p-values. Statistical significance was defined as p<0.05; values near this threshold are noted (e.g., † p=0.051). Discrimination was summarized using the Harrell C-index. Proportional hazards were assessed using standard diagnostics (Schoenfeld residuals).

Associations among immune, spatial, and histopathology variables were evaluated using Spearman correlation (continuous–continuous), η² from type-II ANOVA (categorical–continuous), and Cramér’s V (categorical–categorical). Significance was tested with Pearson correlation tests, Kruskal–Wallis tests, or χ² tests with Monte Carlo estimation for small cell counts. P-values were adjusted using the Benjamini–Hochberg false discovery rate (FDR). For visualization, the top 25 associations by effect size (|r|, η², or V) were displayed as lollipop plots, with significance denoted as ***q<0.001, **q<0.01, *q<0.05, •q<0.10.

All analyses were conducted in R v4.4.0 (survival v3.7-0 and companion packages); figures were generated with ggplot2 and patchwork.

## Data and Code Availability

Code is available at: https://gitlab.com/horlings-lab/youngboost. Requests for sequencing data and pathology images will be reviewed by the NKI IRB and require a material/data transfer agreement (contact: HMH).

## Authors’ Contributions

I.N., S.H., M.J.vd.V., and H.M.H. conceived the project. I.N. collected longitudinal samples and clinical data; S.B. contributed to clinical data collection. A.S., H.B., L.J.B., and P.P. coordinated trial procedures. M.d.M. processed FFPE material, performed immunohistochemistry, isolated RNA, and constructed tissue microarrays. M.L.H.S., N.C.M.V., J.S., M.J.vd.V., and H.M.H. performed centralized pathology review. M.K. and R.S. provided expertise in breast cancer immunology and clinical oncology. E.v.W. advised on statistical analyses. I.H. developed and implemented the multiplex triplex staining protocol. D.P. and R.E. processed multiplex-stained images. A.B. supervised molecular pathology techniques. M.I. and N.d.M. generated and supported analysis of MIBI data. R.H. and S.H. processed MIBI and multiplex IHC images. S.H., H.M.H., and I.N. developed the H&E datasets and the deep learning based cell type classification. I.N. and S.H. analyzed image-derived data. B.A.B. and Y.K. processed and analyzed RNA-seq data. I.N., S.H., and R.H. prepared figures. I.N., R.H., S.H., M.J.vd.V., and H.M.H. wrote the manuscript. M.J.vd.V., H.B., and H.M.H. supervised the project. All authors reviewed and approved the final manuscript.

## Acknowledgments

This work was supported by the Dutch Cancer Foundation (grant KWF-10510). We thank the NKI-AVL Core Facility for Molecular Pathology & Biobanking (CFMPB) for technical support. We are grateful to Alain Fourquet, French project lead, and to the following trial investigators for their contributions: L. Scheijmans (Tilburg), A.H. Westenberg (Arnhem), D. Schinagl (Nijmegen), M. van Hezewijk (Arnhem), M.J.C. van der Sangen (Eindhoven), M.C. Stenfert Kroese (Deventer), J.J. Jobsen (Enschede), J.M. Immink (Delft), M.E. Mast (Den Haag), F.M. Gescher (Den Haag), N. Bijker (Amsterdam), J.W.M. Mens (Rotterdam), W.G.J.M. Smit (Leeuwarden), D.H.F. Rietveld (Amsterdam), J.H. Maduro (UMCG, Groningen), C. Lemansky (Montpellier), I. Lecouillard (Rennes), C. Breton-Callu (Bordeaux), S. Rivera (Villejuif), J. Thariat (Nice), A. Benyoucef (Rouen), A. Labib (Saint Cloud), M. Aumont (Saint Herblain), P. Bontemps (Besançon), C. Le Foll (Lagny), Y. Belkacemi (Créteil), O. Chapet (Lyon), and V. Strnad (Erlangen).

---

**Supplementary Figure 1.**
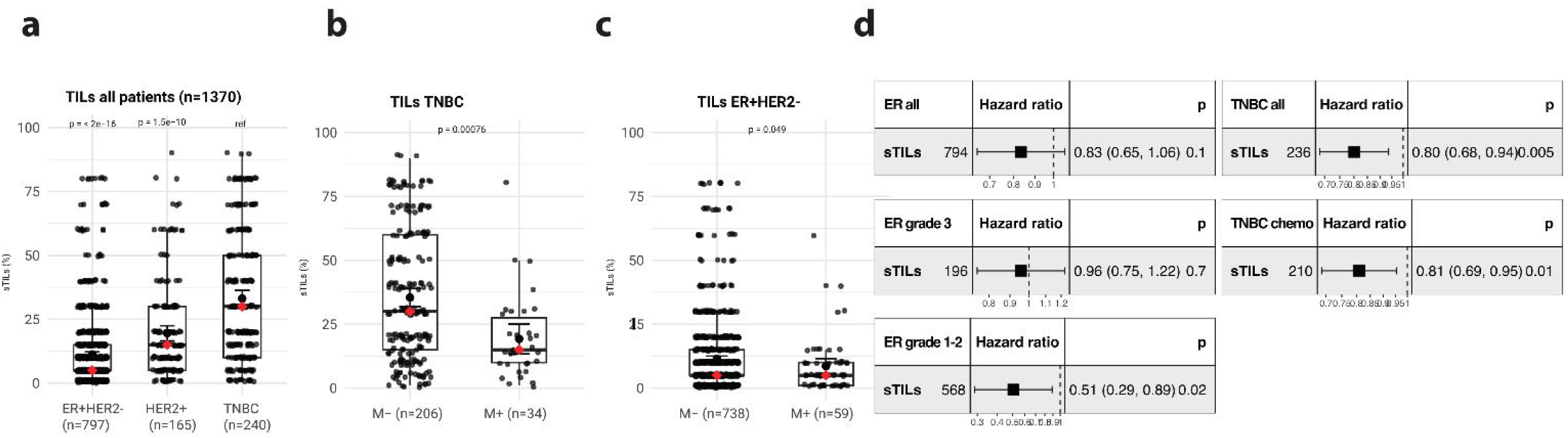
| sTILs and univariate analysis of sTILs in subgroups. a, Levels of sTILs in ER+HER2-, HER2+, and TNBC patients. b, Levels of sTILs in TNBC patients with (M+) and without (M-) subsequent distant metastasis. c, Levels of sTILs in ER+HER2- breast cancer patients with (M+) and without (M-) subsequent distant metastasis. d, Univariate cox models (DMFS) for sTILs in different subgroups of patients; all TNBC patients, TNBC patients who received adjuvant chemo, all ER+HER2- patients, ER+HER2- grade 1-2 patients, grade 3 ER+HER2- patients.

**Supplementary Fig. 2.**
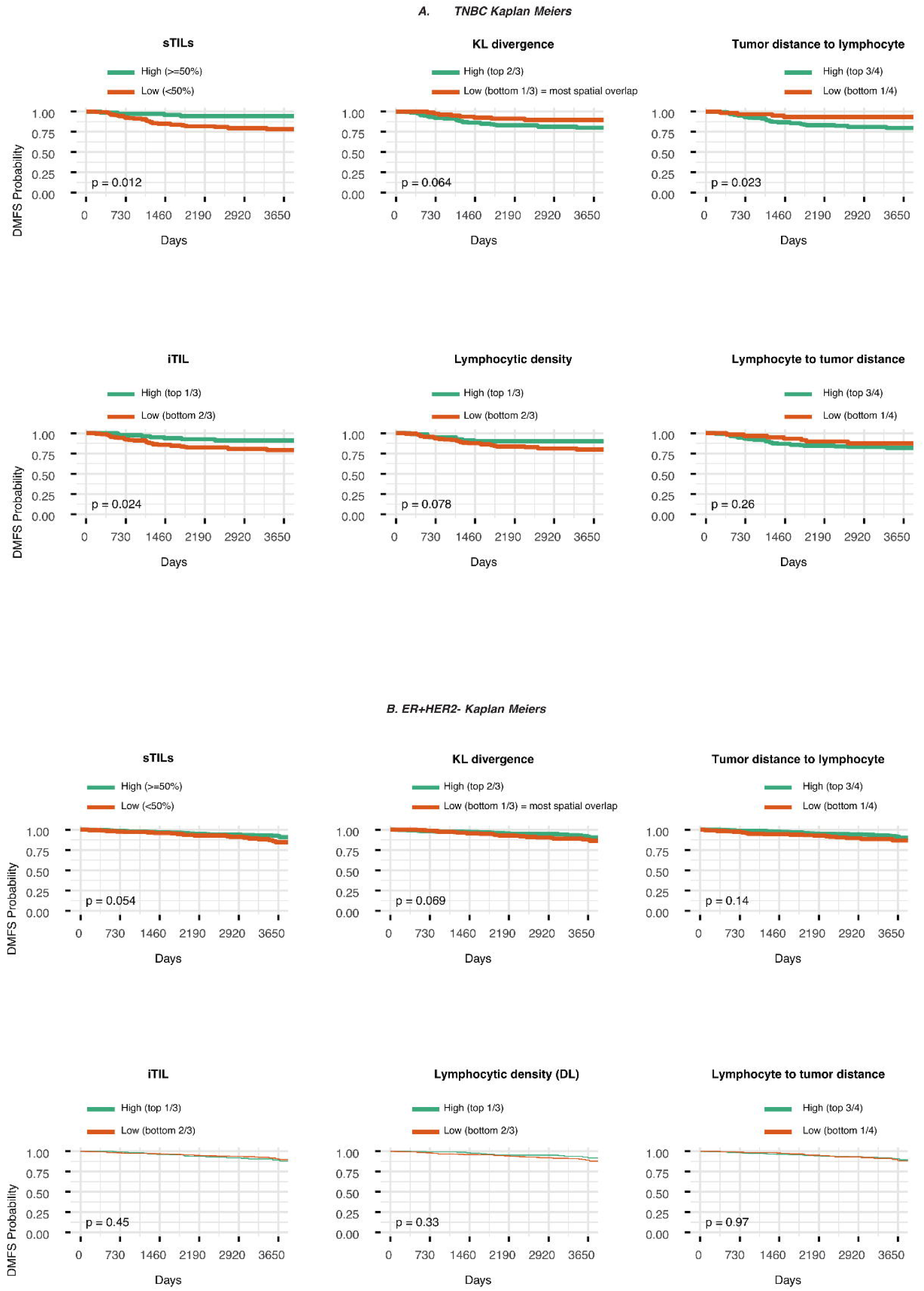
| Kaplan–Meier analyses by subtype. a, TNBC. b, ER⁺/HER2⁻. DMFS curves stratified by six prespecified features: sTILs (TNBC ≥50% vs <50%; ER⁺/HER2⁻ >5% vs ≤5%), iTIL (top tertile vs lower two tertiles), KL divergence (low = bottom tertile vs high = top tertile), model derived lymphocytic densities (top tertile vs lower two tertiles), tumor→lymphocyte distance, and lymphocyte→tumor distance (both low = bottom quartile vs high = upper three quartiles). Time shown in years since diagnosis (x-axis scaled from days). Two-sided log-rank p values indicated. Confidence bands omitted for clarity; censoring ticks shown. Cut-points fixed a priori (Methods). Abbreviations: DMFS, distant metastasis–free survival; sTILs, stromal tumor-infiltrating lymphocytes; iTIL, mixing index; KL, Kullback–Leibler divergence.

**Supplementary figure 3.**
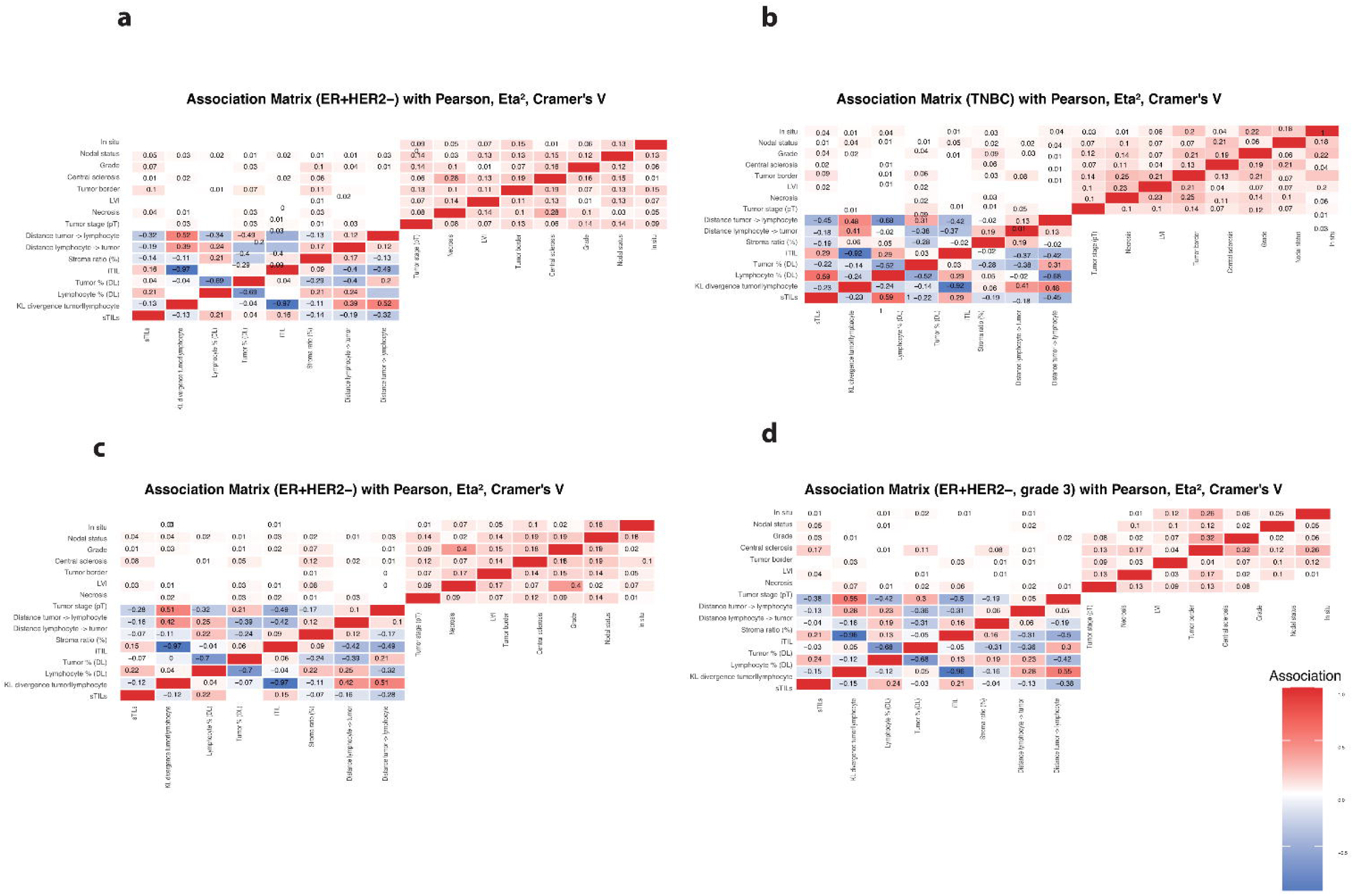
| Association matrices between architecture, immune, and spatial features. a–d, Association matrices for (a) all ER⁺/HER2⁻ tumors, (c) ER⁺/HER2⁻ grade 1–2, (d) ER⁺/HER2⁻ grade 3, and (b) TNBC. Pairwise relationships were quantified using Spearman correlation (continuous–continuous), η² from type-II ANOVA (categorical–continuous), and Cramér’s V (categorical–categorical). Significance was assessed with Pearson correlation tests, Kruskal–Wallis tests, or χ² tests with Monte Carlo estimation for small cell counts. P values were adjusted using the Benjamini–Hochberg false discovery rate (FDR).

**Supplementary Fig. 4.**
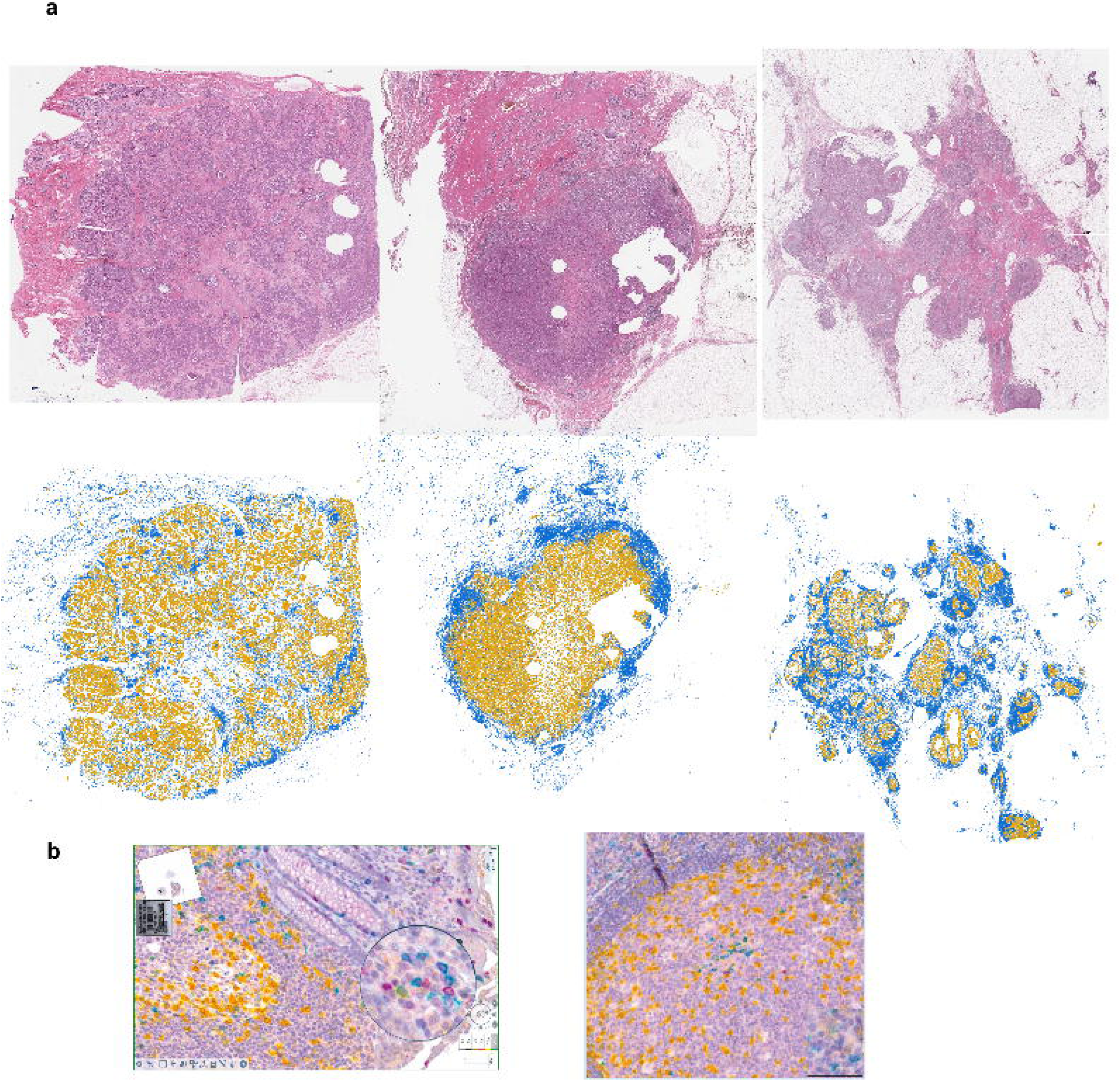
| Example images from H&E and IHC analysis. a, Output from the deep-learning H&E classification pipeline: raw H&E (top) and classified map (bottom), with tumor cells (yellow) and lymphocytes (blue). b, Representative multiplex IHC triple immunodetection.

**Supplementary Fig. 5.**
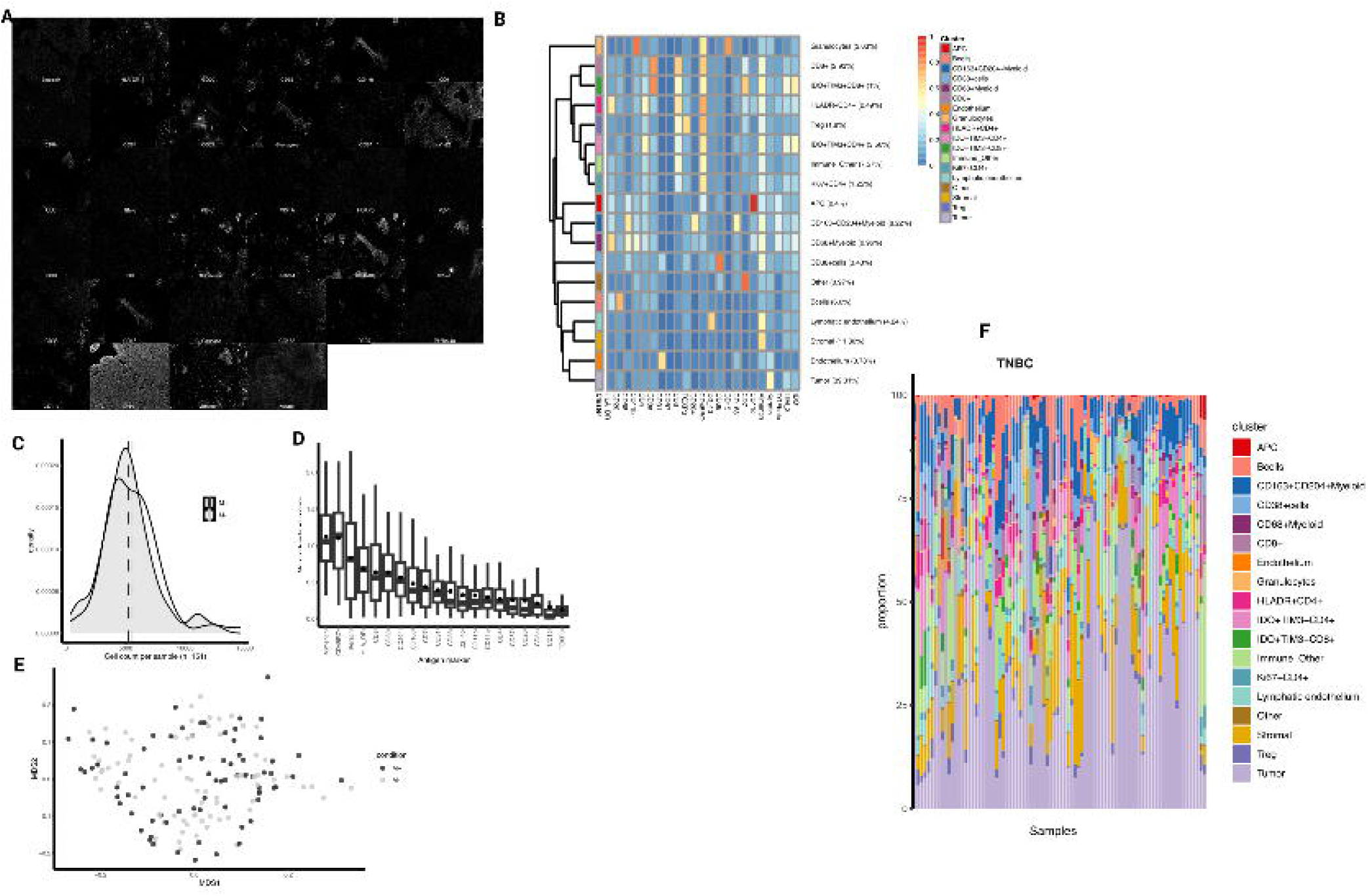
| Stepwise process of phenotyping cells for imaging mass cytometry. a) The image analysis starts with a multi-channel dataset (40 channels, 1x1 µm ‘pixels’) where we measure intensities in all channels. b) Heatmap of the median marker intensities of the lineage markers across 18 cell populations obtained with FlowSOM after the meta clustering step with ConsensusClusterPlus. The color in the heatmap represents the median of the arcsinh, 0-1 transformed marker expression calculated over cells from all the samples, varying from blue for lower expression to red for higher expression. The numbers indicate the actual expression values. The dendrogram on the left represents the hierarchical similarity between the 18 cell clusters (metric: Euclidean distance; linkage: average). Each cluster has a unique color assigned (bar on the left). Values in the brackets next to the cluster numbers indicate the relative size of clusters. c) Density of cells measured for each sample in the IMC dataset. Dotted line indicates median for M+ and M- TNBC patients. d) Non-redundancy scores for each of the 20 lineage markers and all samples in the TNBC dataset. The full points represent the per-sample NR scores, while empty black circles indicate the mean NR scores from all the samples. Markers on the x-axis are sorted according to the decreasing average NRS. e) MDS plot for the M+ and M- TNBC samples. Calculations are based on the median (arcsinh-transformed) marker expression of 20 lineage markers and 12 functional markers across all cells measured for each sample. Distances between samples in the plot approximate the typical change in medians. f) Fractions of cell types across the TNBC cohort.

**Supplementary Table 1.** Comparison of baseline clinical and pathological characteristics between patients with available and evaluable FFPE tumor material (n = 1370) and those without suitable archival FFPE material (n = 1051). Among the 1481 tumor samples retrieved, 1370 were deemed representative and included in the analysis. Categorical variables were compared using Pearson’s chi-squared test, while continuous variables were assessed using the Kruskal–Wallis rank sum test. Values are presented as counts and percentages unless otherwise indicated.

| Characteristic | No FFPE (N = 1051) | FFPE Available (N = 1370) | p-value |
| --- | --- | --- | --- |
| **ER status** |  |  | 0.001 |
| Negative | 192 (18.3%) | 326 (23.8%) |  |
| Positive | 855 (81.7%) | 1043 (76.2%) |  |
| **Age (years)** |  |  | 0.023 |
| Median (IQR) | 44.0 (40.0–47.5) | 45.0 (41.0–48.0) |  |
| **Nodal status** |  |  | 0.775 |
| N0 | 736 (70.6%) | 958 (70.0%) |  |
| N+ | 307 (29.4%) | 410 (30.0%) |  |
| **Menopausal status** |  |  | 0.002 |
| Pre | 895 (87.4%) | 1213 (91.4%) |  |
| Post | 129 (12.6%) | 114 (8.6%) |  |
| **Breast cancer subtype** |  |  | 0.743 |
| ER<sup>+</sup>HER2<sup>-</sup> | 266 (67.7%) | 797 (66.3%) |  |
| HER2<sup>+</sup> | 48 (12.2%) | 165 (13.7%) |  |
| TNBC | 79 (20.1%) | 240 (20.0%) |  |
| **Tumor size** |  |  | 0.004 |
| T1 | 781 (75.1%) | 953 (69.8%) |  |
| T2–3 | 259 (24.9%) | 413 (30.2%) |  |
| **Histological subtype** |  |  | 0.679 |
| NST | 10 (90.9%) | 1252 (91.4%) |  |
| Lobular | 1 (9.1%) | 68 (5.0%) |  |
| Other | 0 (0.0%) | 50 (3.6%) |  |
| **Metastatic progression** |  |  | 0.002 |
| No | 997 (94.9%) | 1256 (91.7%) |  |
| Yes | 54 (5.1%) | 114 (8.3%) |  |

**Abbreviations:** FFPE, Formalin-Fixed Paraffin-Embedded; ER, Estrogen Receptor; IQR, Interquartile Range; HER2, Human Epidermal Growth Factor Receptor 2; TNBC, Triple-Negative Breast Cancer; NST, No Special Type; T1, T2–3, Tumor size categories (TNM classification).

**Table S2.**
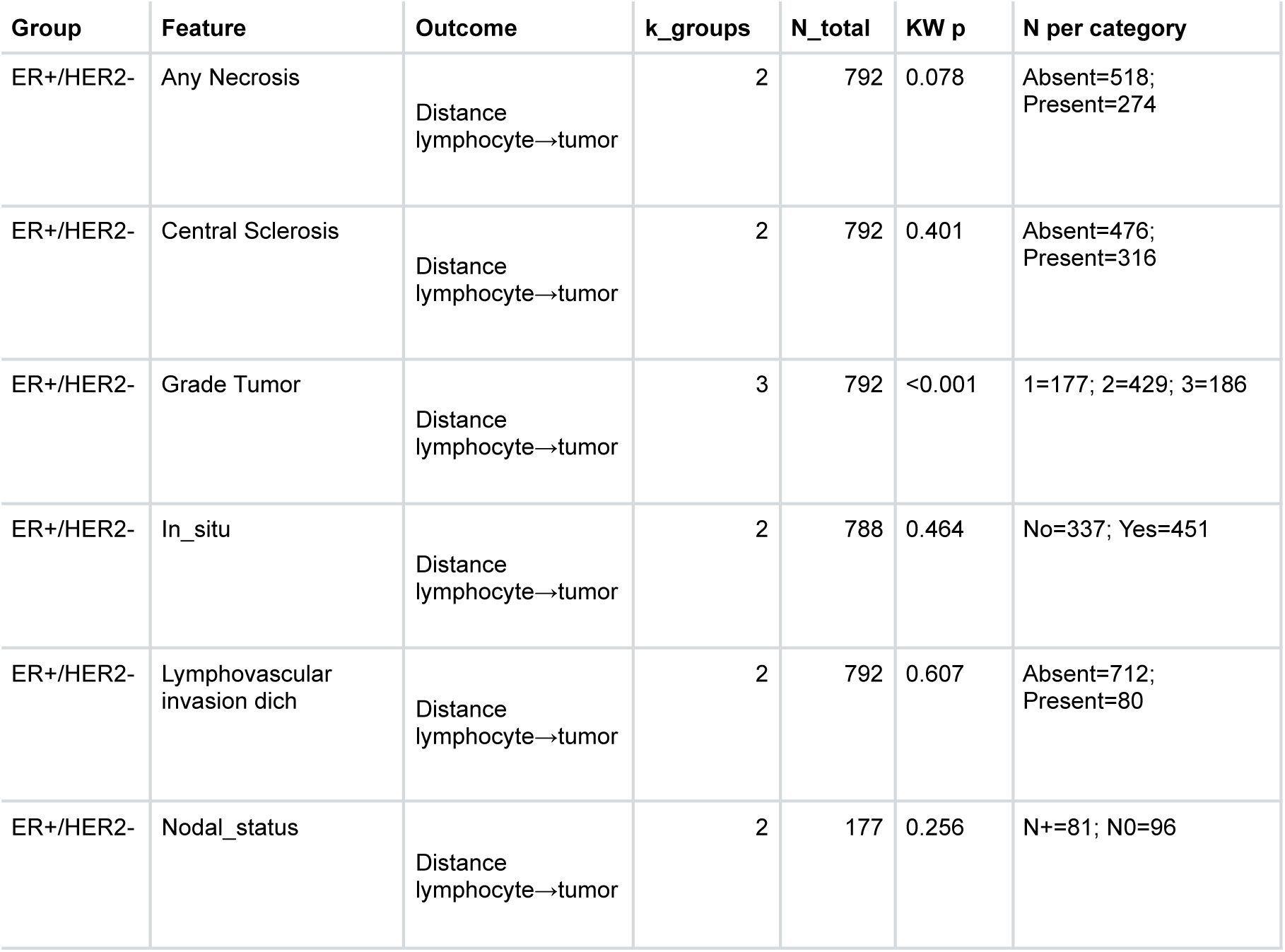
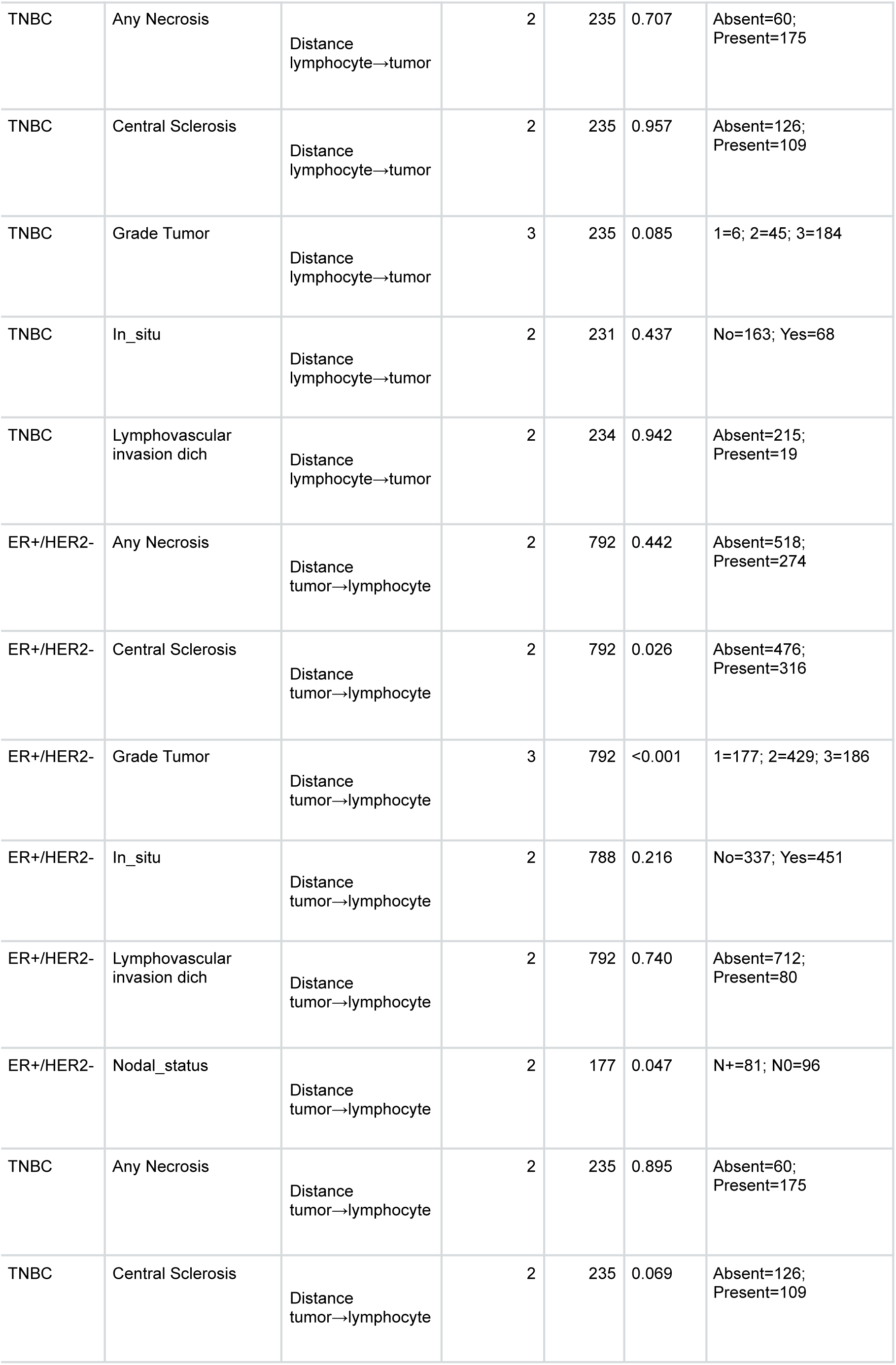
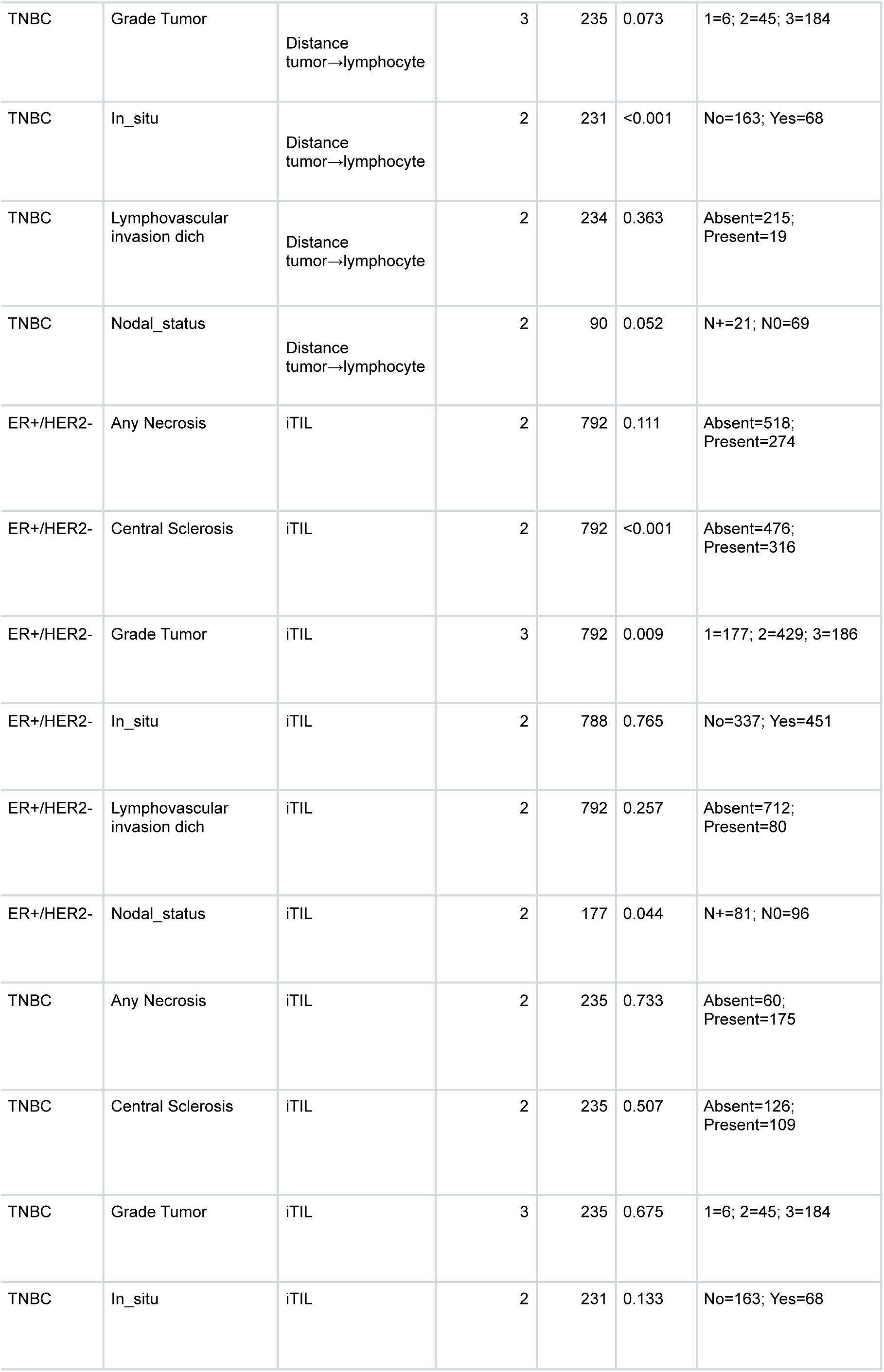
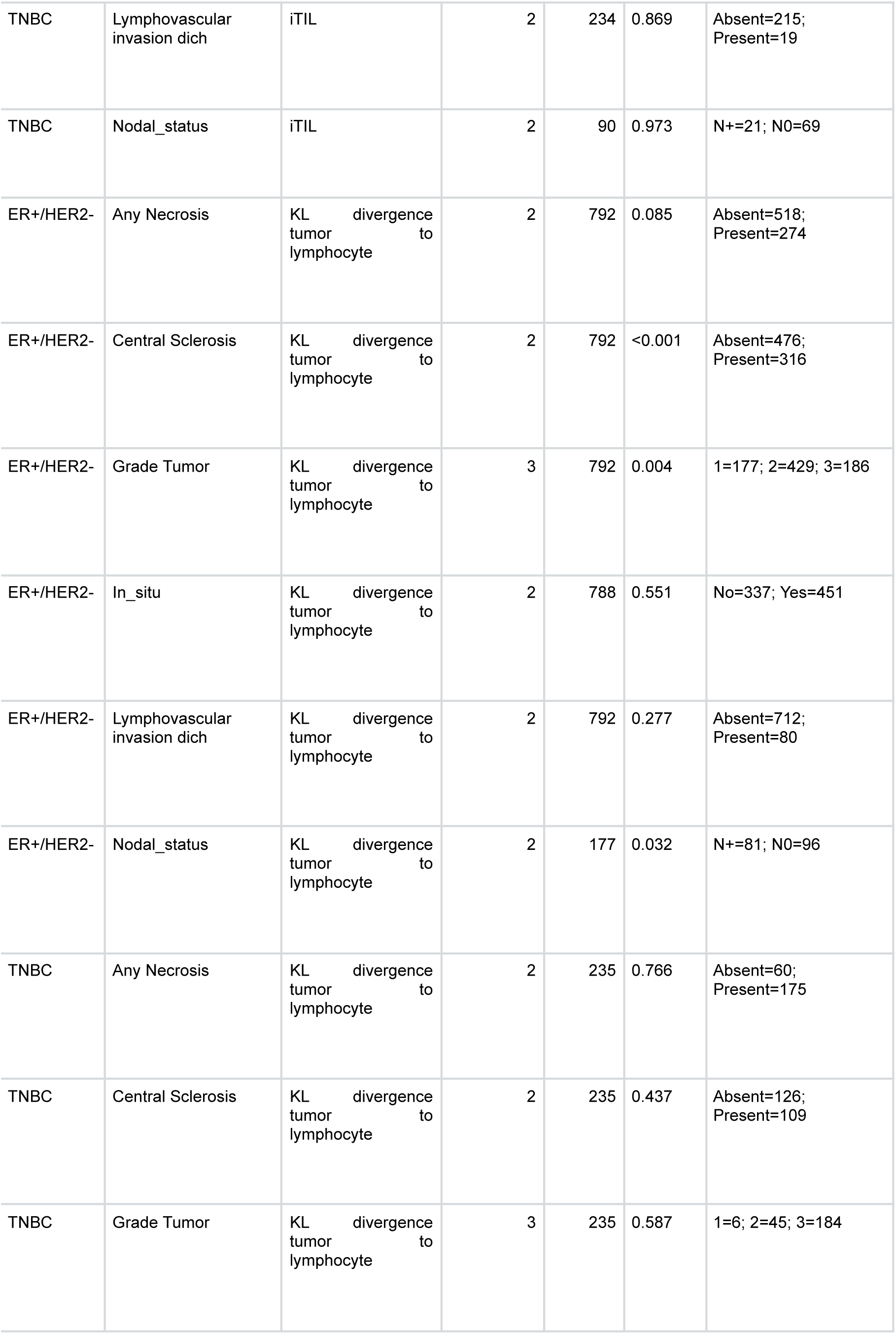
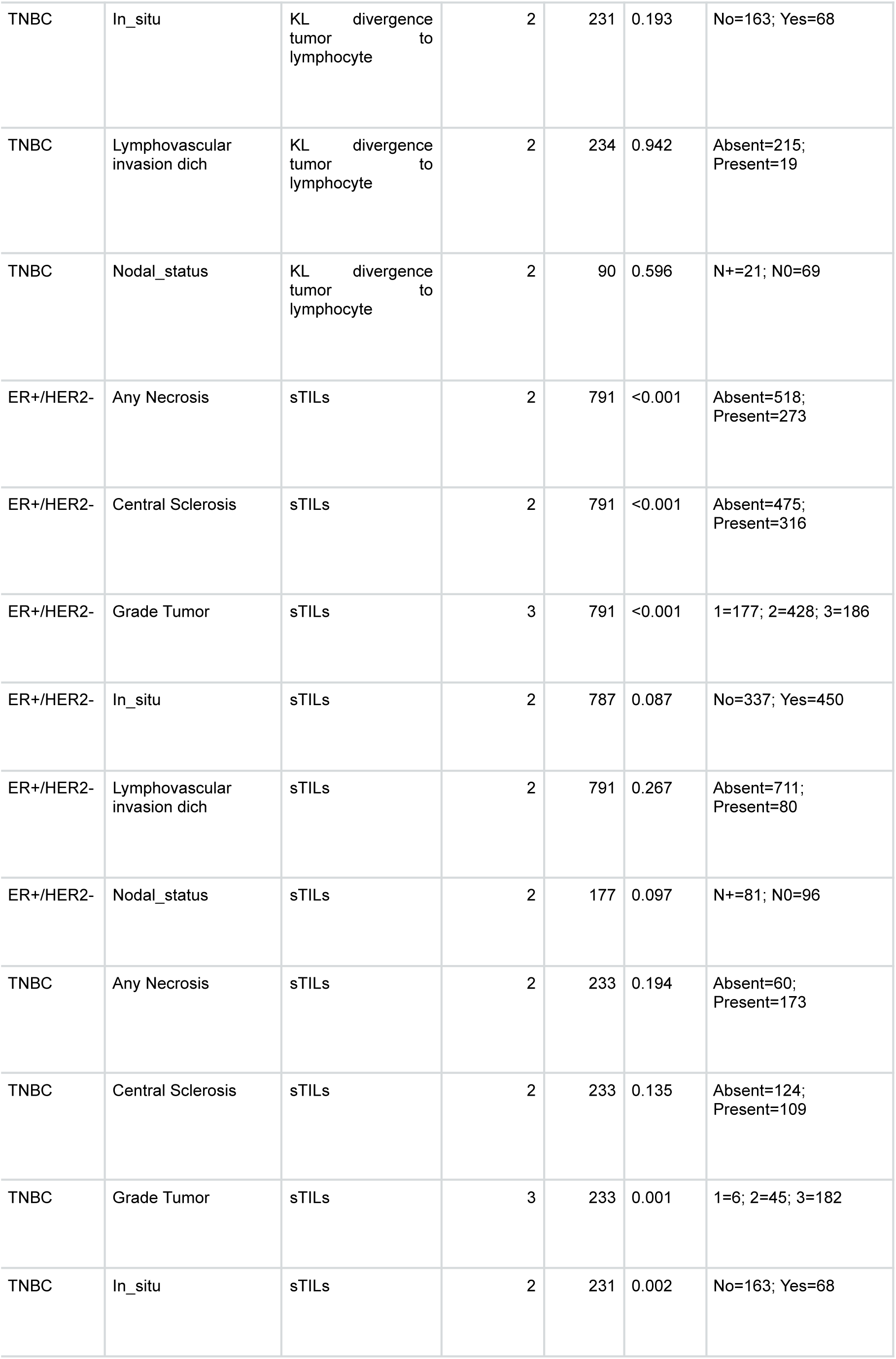
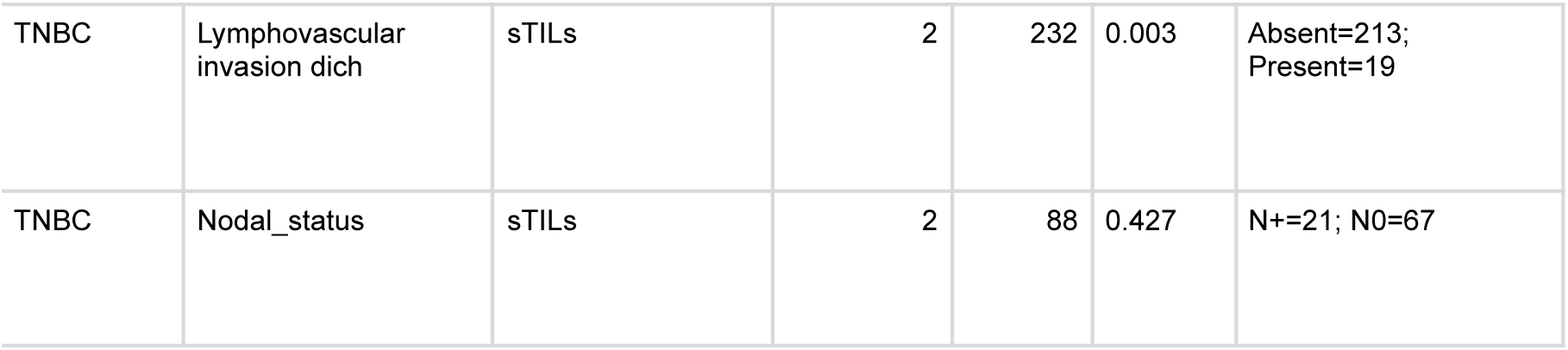
Per-facet Kruskal–Wallis tests of immune metrics across clinicopathologic categories, stratified by subtype (ER⁺/HER2⁻ and TNBC). Each row reports one outcome—stromal TILs (sTILs, %), intratumoral TIL score (iTIL, unitless), lymphocyte→tumor and tumor→lymphocyte nearest-neighbor distances (L→T, T→L, µm), and tumor–lymphocyte KL divergence (unitless)—tested across levels of a feature (e.g., grade, central sclerosis, nodal status, LVI, necrosis, in-situ component). Columns show the number of groups compared (k_groups), total N analyzed (N_total), the Kruskal–Wallis p-value (KW p), and a compact breakdown of N per category used in the test. Only facets with ≥2 categories and non-constant outcome were tested; categories labeled “NA” were excluded. P-values are two-sided and not adjusted across facets. Units: distances in µm; KL is dimensionless; sTILs in %.

**Table S4.** Significant pairwise Wilcoxon comparisons (BH-adjusted) within each facet and subtype. For each outcome (sTILs, iTIL, KL divergence, L→T, T→L) and feature, the table lists the Contrast (second level vs first level), sample sizes (n1, n2) and medians (med1, med2) for the two levels, the BH-adjusted p-value (Adj p-value), the Direction arrow (↑ if median₂>median₁, ↓ if median₂<median₁), and the Median diff (median₂ − median₁) with Units (µm for distances; unitless for KL and iTIL; % for sTILs). Only comparisons with FDR<0.05 are shown. Facets were included only if the outcome varied and each compared category had ≥5 observations; categories labeled “NA” and nodal micro-metastasis were excluded. Interpretation notes: higher KL indicates greater tumor–lymphocyte separation; shorter distances indicate closer spatial proximity.

| Group | Feature | Outcome | Contrast | n1 | n2 | med1 | med2 | Adj p-value | Direction | Median diff | Units |
| --- | --- | --- | --- | --- | --- | --- | --- | --- | --- | --- | --- |
| ER+/HER2- | Grade Tumor | d_lymphocyte_to_tumor | 1 vs 2 | 429 | 177 | 189.771 | 217.183 | <0.001 | ↑ | 27.412 | (μm) |
| ER+/HER2- | Grade Tumor | d_lymphocyte_to_tumor | 1 vs 3 | 186 | 177 | 154.447 | 217.183 | <0.001 | ↑ | 62.737 | (μm) |
| ER+/HER2- | Grade Tumor | d_lymphocyte_to_tumor | 2 vs 3 | 186 | 429 | 154.447 | 189.771 | <0.001 | ↑ | 35.324 | (μm) |
| ER+/HER2- | Central Sclerosis | d_tumor_to_lymphocyte | Absent vs Present | 316 | 476 | 75.221 | 78.852 | 0.026 | ↑ | 3.632 | (μm) |
| ER+/HER2- | Grade Tumor | d_tumor_to_lymphocyte | 1 vs 2 | 429 | 177 | 77.269 | 84.247 | <0.001 | ↑ | 6.978 | (μm) |
| ER+/HER2- | Grade Tumor | d_tumor_to_lymphocyte | 1 vs 3 | 186 | 177 | 71.883 | 84.247 | <0.001 | ↑ | 12.365 | (μm) |
| ER+/HER2- | Grade Tumor | d_tumor_to_lymphocyte | 2 vs 3 | 186 | 429 | 71.883 | 77.269 | 0.012 | ↑ | 5.386 | (μm) |
| ER+/HER2- | Nodal_status | d_tumor_to_lymphocyte | N+ vs N0 | 96 | 81 | 74.354 | 79.393 | 0.047 | ↑ | 5.040 | (μm) |
| TNBC | In_situ | d_tumor_to_lymphocyte | No vs Yes | 68 | 163 | 74.821 | 62.662 | <0.001 | ↓ | -12.160 | (μm) |
| ER+/HER2- | Central Sclerosis | itil | Absent vs Present | 316 | 476 | 0.092 | 0.020 | <0.001 | ↓ | -0.073 | (score) |
| ER+/HER2- | Grade Tumor | itil | 1 vs 2 | 429 | 177 | 0.072 | -0.004 | 0.007 | ↓ | -0.076 | (score) |
| ER+/HER2- | Grade Tumor | itil | 1 vs 3 | 186 | 177 | 0.054 | -0.004 | 0.043 | ↓ | -0.058 | (score) |
| ER+/HER2- | Nodal_status | itil | N+ vs N0 | 96 | 81 | 0.152 | 0.048 | 0.044 | ↓ | -0.104 | (score) |
| ER+/HER2- | Central Sclerosis | kl_tl | Absent vs Present | 316 | 476 | 0.806 | 0.886 | <0.001 | ↑ | 0.080 | (unitless) |
| ER+/HER2- | Grade Tumor | kl_tl | 1 vs 2 | 429 | 177 | 0.835 | 0.912 | 0.003 | ↑ | 0.078 | (unitless) |
| ER+/HER2- | Grade Tumor | kl_tl | 1 vs 3 | 186 | 177 | 0.854 | 0.912 | 0.033 | ↑ | 0.058 | (unitless) |
| ER+/HER2- | Nodal_status | kl_tl | N+ vs N0 | 96 | 81 | 0.719 | 0.851 | 0.032 | ↑ | 0.132 | (unitless) |
| ER+/HER2- | Any Necrosis | sTILs | Absent vs Present | 273 | 518 | 1.000 | 0.500 | <0.001 | ↓ | -5.00 | % |
| ER+/HER2- | Central Sclerosis | sTILs | Absent vs Present | 316 | 475 | 1.000 | 0.500 | <0.001 | ↓ | -5.00 | % |
| ER+/HER2- | Grade Tumor | sTILs | 1 vs 3 | 186 | 177 | 1.000 | 0.500 | <0.001 | ↓ | -5.00 | % |
| ER+/HER2- | Grade Tumor | sTILs | 2 vs 3 | 186 | 428 | 1.000 | 0.500 | <0.001 | ↓ | -5.00 | % |
| TNBC | Grade Tumor | sTILs | 1 vs 3 | 182 | 6 | 3.000 | 0.750 | 0.012 | ↓ | -22.50 | % |
| TNBC | Grade Tumor | sTILs | 2 vs 3 | 182 | 45 | 3.000 | 2.000 | 0.024 | ↓ | -10.00 | % |
| TNBC | In_situ | sTILs | No vs Yes | 68 | 163 | 1.750 | 3.000 | 0.002 | ↑ | 12.50 | % |
| TNBC | Lymphovascular invasion dich | sTILs | Absent vs Present | 19 | 213 | 1.000 | 3.000 | 0.003 | ↑ | 20.00 | % |

**Table S4.** Significant architecture-immune associations in grade-stratified ER⁺/HER2⁻ disease. (BH-adjusted pairwise Wilcoxon tests within each facet; only FDR < 0.05 shown)

| Feature | Outcome | Contrast | Adj p-value |
| --- | --- | --- | --- |
| **Grade 1-2** |  |  |  |
| **Grade Tumor** | Lymphocyte→Tumor distance | 2 vs 1 | **<0.001** |
|  | Tumor→Lymphocyte distance | 2 vs 1 | **<0.001** |
|  | iTIL | 2 vs 1 | **0.002** |
|  | KL divergence | 2 vs 1 | **<0.001** |
| **Central Sclerosis** | Tumor→Lymphocyte distance | Present vs Absent | **0.023** |
|  | iTIL | Present vs Absent | **<0.001** |
|  | KL divergence | Present vs Absent | **<0.001** |
|  | sTILs | Present vs Absent | **<0.001** |
| **Nodal status** | Tumor→Lymphocyte distance | N0 vs N+ | **0.048** |
|  | KL divergence | N0 vs N+ | **0.039** |
| **Any Necrosis** | sTILs | Present vs Absent | **<0.001** |
| **In-situ** | sTILs | Yes vs No | **0.044** |
| **Grade 3** |  |  |  |
| **Lymphovascular invasion** | iTIL | Present vs Absent | **0.024** |
|  | KL divergence | Present vs Absent | **0.022** |
| **Any Necrosis** | sTILs | Present vs Absent | **<0.001** |

*Abbreviations:* iTIL, intratumoral TIL score (unitless); sTILs, stromal TILs (%); KL, tumor-lymphocyte KL divergence (unitless). Distances are nearest-neighbor in $\mu\text{m}$ .

## REFERENCES

1. Park, J. H. et al. Prognostic value of tumor-infiltrating lymphocytes in patients with early-stage triple-negative breast cancers (TNBC) who did not receive adjuvant chemotherapy. Ann. Oncol. 30, 1941–1949 (2019).

2. Loi, S. et al. Tumor-Infiltrating Lymphocytes and Prognosis: A Pooled Individual Patient Analysis of Early-Stage Triple-Negative Breast Cancers. J. Clin. Oncol. 37, 559–569 (2019).

3. Denkert, C. et al. Tumour-infiltrating lymphocytes and prognosis in different subtypes of breast cancer: a pooled analysis of 3771 patients treated with neoadjuvant therapy. Lancet Oncol. 19, 40–50 (2018).

4. Kos, Z. et al. Pitfalls in assessing stromal tumor infiltrating lymphocytes (sTILs) in breast cancer. NPJ Breast Cancer 6, 17 (2020).

5. Salgado, R. et al. The evaluation of tumor-infiltrating lymphocytes (TILs) in breast cancer: recommendations by an International TILs Working Group 2014. Ann. Oncol. 26, 259–271 (2015).

6. Fridman, W. H., Pagès, F., Sautès-Fridman, C. & Galon, J. The immune contexture in human tumours: impact on clinical outcome. Nature Reviews Cancer 12, 298–306 (2012).

7. Joyce, J. A. & Fearon, D. T. T cell exclusion, immune privilege, and the tumor microenvironment. Science (2015) doi:10.1126/science.aaa6204.

8. Gruosso, T. et al. Spatially distinct tumor immune microenvironments stratify triple-negative breast cancers. J. Clin. Invest. 129, 1785–1800 (2019).

9. Hammerl, D. et al. Spatial immunophenotypes predict response to anti-PD1 treatment and capture distinct paths of T cell evasion in triple negative breast cancer. Nat. Commun. 12, 5668 (2021).

10. Cardoso, F. et al. Pembrolizumab and chemotherapy in high-risk, early-stage, ER/HER2 breast cancer: a randomized phase 3 trial. Nat Med 31, 442–448 (2025).

11. Loi, S. et al. Neoadjuvant nivolumab and chemotherapy in early estrogen receptor-positive breast cancer: a randomized phase 3 trial. Nat Med 31, 433–441 (2025).

12. Colpaert, C., Vermeulen, P., Van Marck, E. & Dirix, L. The presence of a fibrotic focus is an independent predictor of early metastasis in lymph node-negative breast cancer patients. The American journal of surgical pathology 25, (2001).

13. Kreike, B. et al. Gene expression profiling and histopathological characterization of triple-negative/basal-like breast carcinomas. Breast cancer research : BCR 9, (2007).

14. Mariathasan, S. et al. TGFβ attenuates tumour response to PD-L1 blockade by contributing to exclusion of T cells. Nature 554, 544–548 (2018).

15. Accessories to the Crime: Functions of Cells Recruited to the Tumor Microenvironment. Cancer Cell 21, 309–322 (2012).

16. Quail, D. F. & Joyce, J. A. Microenvironmental regulation of tumor progression and metastasis. Nature medicine 19, (2013).

17. Turley, S. J., Cremasco, V. & Astarita, J. L. Immunological hallmarks of stromal cells in the tumour microenvironment. Nature reviews. Immunology 15, (2015).

18. Kalluri, R. The biology and function of fibroblasts in cancer. Nature Reviews Cancer 16, 582–598 (2016).

19. Costa, A. et al. Fibroblast Heterogeneity and Immunosuppressive Environment in Human Breast Cancer. Cancer Cell 33, 463–479.e10 (2018).

20. Risom, T. et al. Transition to invasive breast cancer is associated with progressive changes in the structure and composition of tumor stroma. Cell 185, 299–310.e18 (2022).

21. The evolving tumor microenvironment: From cancer initiation to metastatic outgrowth. Cancer Cell 41, 374–403 (2023).

22. Bagaev, A. et al. Conserved pan-cancer microenvironment subtypes predict response to immunotherapy. Cancer cell 39, (2021).

23. Virassamy, B. et al. Intratumoral CD8+ T cells with a tissue-resident memory phenotype mediate local immunity and immune checkpoint responses in breast cancer. Cancer Cell 41, 585–601.e8 (2023).

24. Savas, P. et al. Single-cell profiling of breast cancer T cells reveals a tissue-resident memory subset associated with improved prognosis. Nat. Med. 24, 986–993 (2018).

25. Beavis, P. A., Mackay, L. K., Neeson, P. J. & Loi, S. Tissue-resident memory T cells in breast cancer control and immunotherapy responses. Nat. Rev. Clin. Oncol. (2020).

26. Schumacher, T. N. & Thommen, D. S. Tertiary lymphoid structures in cancer. Science (2022) doi:10.1126/science.abf9419.

27. Bosma, S. C. J. et al. A case-control study to identify molecular risk factors for local recurrence in young breast cancer patients. Radiother. Oncol. 156, 127–135 (2021).

28. Brouwers, P. J. A. M. et al. Predictors for poor cosmetic outcome in patients with early stage breast cancer treated with breast conserving therapy: Results of the Young boost trial. Radiother. Oncol. 128, 434–441 (2018).

29. Early breast cancer: ESMO Clinical Practice Guideline for diagnosis, treatment and follow-up. Annals of Oncology 35, 159–182 (2024).

30. slidescore.com. https://slidescore.com.

31. Hasebe, T. et al. Fibrotic focus in invasive ductal carcinoma: an indicator of high tumor aggressiveness. Japanese journal of cancer research : Gann 87, (1996).

32. Maiorano, E. et al. Prognostic and predictive impact of central necrosis and fibrosis in early breast cancer: results from two International Breast Cancer Study Group randomized trials of chemoendocrine adjuvant therapy. Breast cancer research and treatment 121, (2010).

33. Cakir, A., Gonul, I. I. & Uluoglu, O. A comprehensive morphological study for basal-like breast carcinomas with comparison to nonbasal-like carcinomas. Diagnostic pathology 7, (2012).

34. Livasy, C. A. et al. Phenotypic evaluation of the basal-like subtype of invasive breast carcinoma. Modern pathology : an official journal of the United States and Canadian Academy of Pathology, Inc 19, (2006).

35. den Eynden GG, V., et al. A fibrotic focus is a prognostic factor and a surrogate marker for hypoxia and (lymph)angiogenesis in breast cancer: review of the literature and proposal on the criteria of evaluation. Histopathology 51, (2007).

36. WHO Classification of Tumours Editorial Board. WHO Classification of Breast Tumours: WHO Classification of Tumours, Volume 2. (International Agency for Research on Cancer, 2019).

37. Van der Leun, A. M. & Thommen, D. S. CD8+ T cell states in human cancer: insights from single-cell analysis. Nat. Rev. (2020).

38. Corgnac, S., Boutet, M., Kfoury, M., Naltet, C. & Mami-Chouaib, F. The Emerging Role of CD8 Tissue Resident Memory T (T) Cells in Antitumor Immunity: A Unique Functional Contribution of the CD103 Integrin. Front Immunol 9, 1904 (2018).

39. Yenyuwadee, S., Lopez, J. L. S.-T., Shah, R., Rosato, P. C. & Boussiotis, V. A. The evolving role of tissue-resident memory T cells in infections and cancer. Science Advances (2022) doi:10.1126/sciadv.abo5871.

40. Ahmed, F. S. et al. PD-L1 protein expression on both tumor cells and macrophages are associated with response to neoadjuvant durvalumab with chemotherapy in triple-negative breast cancer. Clin. Cancer Res. 26, 5456–5461 (2020).

41. Ijsselsteijn, M. E., van der Breggen, R., Farina Sarasqueta, A., Koning, F. & de Miranda, N. F. C. C. A 40-Marker Panel for High Dimensional Characterization of Cancer Immune Microenvironments by Imaging Mass Cytometry. Front. Immunol. 10, 2534 (2019).

42. Nowicka, M. et al. CyTOF workflow: differential discovery in high-throughput high-dimensional cytometry datasets. F1000Res. 6, 748 (2017).

43. Chen, Z., Soifer, I., Hilton, H., Keren, L. & Jojic, V. Modeling Multiplexed Images with Spatial-LDA Reveals Novel Tissue Microenvironments. J. Comput. Biol. 27, 1204–1218 (2020).

44. Geurts, V. C. M., et al. Tumor-Infiltrating Lymphocytes in Patients With Stage I Triple-Negative Breast Cancer Untreated With Chemotherapy. JAMA Oncol 10, 1077–1086 (2024).

45. de Jong, V. M. T., et al. Prognostic Value of Stromal Tumor-Infiltrating Lymphocytes in Young, Node-Negative, Triple-Negative Breast Cancer Patients Who Did Not Receive (neo)Adjuvant Systemic Therapy. J. Clin. Oncol. 40, 2361–2374 (2022).

46. Website. https://www.jstor.org/stable/2236703?seq=1.

47. van ’t Veer, L. J., et al. Gene expression profiling predicts clinical outcome of breast cancer. Nature 415, 530–536 (2002).

48. Li, Y., Wang, C., Huang, T., Yu, X. & Tian, B. The role of cancer-associated fibroblasts in breast cancer metastasis. Front. Oncol. 13, 1194835 (2023).

49. Fan, G., et al. TSPAN8+ myofibroblastic cancer-associated fibroblasts promote chemoresistance in patients with breast cancer. Sci. Transl. Med. 16, eadj5705 (2024).

50. Elston, C. W. & Ellis, I. O. Pathological prognostic factors in breast cancer. I. The value of histological grade in breast cancer: experience from a large study with long-term follow-up. Histopathology 19, 403–410 (1991).

51. de Boo, L. W., et al. Prognostic value of histopathologic traits independent of stromal tumor-infiltrating lymphocyte levels in chemotherapy-naïve patients with triple-negative breast cancer. ESMO Open 9, 102923 (2024).

52. Silverberg, S. G., Chitale, A. R. & Levitt, S. H. Prognostic significance of tumor margins in mammary carcinoma. Arch. Surg. 102, 450–454 (1971).

53. Thike, A. A., et al. Triple-negative breast cancer: clinicopathological characteristics and relationship with basal-like breast cancer. Mod. Pathol. 23, 123–133 (2010).

54. Colpaert, C. G., et al. The presence of a fibrotic focus in invasive breast carcinoma correlates with the expression of carbonic anhydrase IX and is a marker of hypoxia and poor prognosis. Breast Cancer Res. Treat. 81, 137–147 (2003).

55 Colpaert, C. et al. Intratumoral hypoxia resulting in the presence of a fibrotic focus is an independent predictor of early distant relapse in lymph node-negative breast cancer patients: Fibrotic focus in breast cancer. Histopathology 39, 416–425 (2001).

56 Gilchrist, K. W., Gray, R., Fowble, B., Tormey, D. C. & Taylor, S. G., 4th. Tumor necrosis is a prognostic predictor for early recurrence and death in lymph node-positive breast cancer: a 10-year follow-up study of 728 Eastern Cooperative Oncology Group patients. J. Clin. Oncol. 11, 1929–1935 (1993).

57. Kramer, C. J. H., et al. The prognostic value of tumour-stroma ratio in primary breast cancer with special attention to triple-negative tumours: a review. Breast Cancer Res. Treat. 173, 55–64 (2019).

58. Moorman, A. M., Vink, R., Heijmans, H. J., van der Palen, J. & Kouwenhoven, E. A. The prognostic value of tumour-stroma ratio in triple-negative breast cancer. Eur. J. Surg. Oncol. 38, 307–313 (2012).

59. Buisseret, L., et al. Reliability of tumor-infiltrating lymphocyte and tertiary lymphoid structure assessment in human breast cancer. Mod. Pathol. 30, 1204–1212 (2017).

60. Hinton, G., Vinyals, O. & Dean, J. Distilling the Knowledge in a Neural Network. arXiv [stat.ML] (2015).

61. Weigert, M., Schmidt, U., Haase, R., Sugawara, K. & Myers, E. Star-convex polyhedra for 3D object detection and segmentation in microscopy. Proc. IEEE Workshop Appl. Comput. Vis. 3655–3662 (2019).

62. Nik-Zainal, S., et al. Landscape of somatic mutations in 560 breast cancer whole-genome sequences. Nature 534, 47–54 (2016).

63 Bottai, G. et al. Abstract P2-04-16: CD103+/PD-1+ T-cells identify a subset of triple-negative breast cancer eligible for targeted checkpoint inhibition. in Poster Session Abstracts (American Association for Cancer Research, 2017). doi:10.1158/1538-7445.sabcs16-p2-04-16.

64. Wang, L., et al. PD-L1-expressing tumor-associated macrophages are immunostimulatory and associate with good clinical outcome in human breast cancer. Cell Rep. Med. 5, 101420 (2024).

65. Suzuki, K., et al. Histological spatial analysis on the induction of PD-L1+ macrophages by CD8+ T cells at the marginal microenvironment of triple-negative breast cancer. Breast Cancer 30, 1094–1104 (2023).

66. Hudeček, J., et al. Application of a risk-management framework for integration of stromal tumor-infiltrating lymphocytes in clinical trials. NPJ Breast Cancer 6, 15 (2020).

67. Bankhead, P., et al. QuPath: Open source software for digital pathology image analysis. Sci. Rep. 7, 16878 (2017).

68. Schindelin, J., et al. Fiji: an open-source platform for biological-image analysis. Nat. Methods 9, 676–682 (2012).

69. Schmidt, U., Weigert, M., Broaddus, C. & Myers, G. Cell detection with star-convex polygons. in Medical Image Computing and Computer Assisted Intervention – MICCAI 2018 265–273 (Springer International Publishing, Cham, 2018).

70. Haase, R., et al. CLIJ: GPU-accelerated image processing for everyone. Nat. Methods 17, 5–6 (2020).

71. Van Gassen, S., et al. FlowSOM: Using self-organizing maps for visualization and interpretation of cytometry data. Cytometry A 87, 636–645 (2015).

72. Wilkerson, M. D. & Hayes, D. N. ConsensusClusterPlus: a class discovery tool with confidence assessments and item tracking. Bioinformatics 26, 1572–1573 (2010).

73. Trapnell, C., Pachter, L. & Salzberg, S. L. TopHat: discovering splice junctions with RNA-Seq. Bioinformatics 25, 1105–1111 (2009).

74. Anders, S., Pyl, P. T. & Huber, W. HTSeq--a Python framework to work with high-throughput sequencing data. Bioinformatics 31, 166–169 (2015).

75. Rooney, M. S., Shukla, S. A., Wu, C. J., Getz, G. & Hacohen, N. Molecular and genetic properties of tumors associated with local immune cytolytic activity. Cell 160, 48–61 (2015).

76. Ayers, M., et al. IFN-γ-related mRNA profile predicts clinical response to PD-1 blockade. J. Clin. Invest. 127, 2930–2940 (2017).

77. Danaher, P., et al. Gene expression markers of Tumor Infiltrating Leukocytes. J Immunother Cancer 5, 18 (2017).

